# Deep cervical lymph nodes of patients with multiple sclerosis show dysregulated B cells in the presence of Epstein-Barr virus

**DOI:** 10.1101/2023.10.22.23297386

**Authors:** Joona Sarkkinen, Dawit Yohannes, Nea Kreivi, Pia Dürnsteiner, Jani Huuhtanen, Kirsten Nowlan, Goran Kurdo, Riikka Linden, Mika Saarela, Pentti J Tienari, Eliisa Kekäläinen, Maria Perdomo, Sini M Laakso

## Abstract

Despite the recognized role of Epstein-Barr virus (EBV) in predisposing to multiple sclerosis (MS) and the effectiveness of B cell-depleting therapies in MS, the mechanism of autoimmunity remains elusive. Using fine needle aspirations, we investigated deep cervical lymph nodes (dcLNs), the primary site of the adaptive immune response against EBV, in newly diagnosed untreated MS patients and healthy controls. We characterized the immune landscape of dcLNs with scRNAseq and CITE-seq and observed increased memory B cell proportions and reduced germinal center (GC) B cells with decreased clonality in patients with MS compared to healthy controls. In the patient with an active MS relapse, we detected elevated plasmablasts, reduced GC B cells, and clonally expanded memory CD8 T cells targeting EBV in the dcLN. These findings, along with increased EBV DNA detection in dcLNs and viral loads in patient saliva, support B cell dysregulation as a key mechanism in MS pathogenesis.

## INTRODUCTION

Multiple sclerosis (MS) is the most common neurological autoimmune disease affecting 2.8 million individuals worldwide and has an increasing prevalence (*1*). Demyelinating inflammatory lesions of the central nervous system (CNS) are the cause of clinical attacks in relapsing-remitting MS (RRMS), which presents in 90% of patients, and migration of lymphocytes to the CNS is the hallmark of the disease pathology (*2*). Despite research efforts, the exact mechanisms that lead to MS remain unknown.

Several environmental and genetic risk factors that predispose to MS have been observed, of which Epstein Barr Virus (EBV) infection in early adulthood is by far the most significant. EBV infection is characterized by the proliferation and transformation of infected B cells and the subsequent proliferation of CD8+ T cells targeting EBV. After primary infection, EBV persists lifelong in a small subset of B cells (*3*). Close to 100% of MS patients are EBV seropositive (*4*, *5*), and recently, Bjornevik et al showed that EBV infection in early adulthood raises the risk for MS by 32-fold (*6*). These epidemiological findings were limited only to EBV, and seropositivity to other herpesviruses such as cytomegalovirus (CMV) does not raise the risk for MS.

Antibodies recognizing EBV may cross-react with self-antigens (*7*). Recently, clonally expanded intrathecal plasmablasts (PBs) were found to produce antibodies that bind to EBV transcription factor EBNA1 but also to glial cell adhesion molecule (GlialCAM). In addition, EBNA1 immunization of experimental autoimmune encephalitis (EAE) mice exacerbated the EAE by increasing autoreactive B and T cell responses. (*8*) EBV-targeting cytotoxic CD8+ T cells have also been detected in autopsy samples of demyelinating brain lesions of MS patients. (*9*, *10*)

The adaptive immune response against EBV occurs most importantly in the deep cervical lymph nodes (dcLN) (*11*), which drain CNS lymphatics carrying neuronal autoantigens also in MS patients (*12–14*). Interestingly, dcLNs are enlarged in MS patients compared to healthy individuals (*15*), and clonally expanded B cells extracted from the demyelinating lesions in MS patients commonly derive from the dcLNs (*16*).

Therefore, it is plausible that dcLN could be the original site where MS pathogenesis is initiated.

These findings, together with the high efficacy of B cell-depleting therapies in treating RRMS, have moved the MS research focus from T cells towards B cells and T-B cell crosstalk. B cell responses are refined in germinal centers (GCs), where B cell somatic hypermutations (SHMs) and class switching take place. GCs are typically found in secondary lymphoid tissues, such as LNs. Ectopic GCs, which develop in response to chronic inflammation (*17*), have also been found on the meninges of MS patients possibly contributing to B cell autoreactivity in MS (*16*, *18*). The GC reaction is regulated by Tfr cells, whereas Tfh cells supply aid to developing B cells, which differentiate into memory B cells (MBCs) and high-affinity antibody (ab) producing PBs and plasma cells (PCs) (*19*). The ratio of follicular T helper cells (Tfh) to follicular regulator cells (Tfr) in the circulation has been reported to be increased in MS and to correlate with intrathecal IgG production, which suggests dysregulation of GC reactions (*20*).

We hypothesized that EBV infection disrupts B cell homeostasis and the GC microenvironment in the dcLNs of MS patients, creating the ground for B cell-mediated autoimmunity. Using fine needle aspirations (FNAs) of the dcLNs (*21*) in newly diagnosed MS patients and healthy controls, we investigated the immunological landscape of dcLNs at single-cell resolution focusing on the GC reaction. We show, for the first time in an *ex vivo* analysis of MS patients, an intranodal expansion of memory B cells and diminished GC B cell proportion and clonality, paralleled by EBV detection.

Our results reveal altered cellular composition of dcLNs in newly diagnosed MS patients, providing insights into the role of EBV in disease pathogenesis and to the efficacy of B cell depleting therapies in RRMS.

## RESULTS

### Investigation of deep cervical lymph nodes accessed by fine needle aspirations

Ultrasound-guided FNAs of dcLNs followed by isolation of viable single cells and immediate processing for 5’ single-cell RNA sequencing including T and B cell receptor sequencing were performed on 7 MS patients (mean age 36 years, 4/9 females) and 6 healthy controls (mean age 28 years, 3/5 females, Fig. 1A). The width and height of dcLNs, measured with ultrasound, did not differ between MS patients and controls (median 0.53 vs 0.49 cm, t-test p = 0.7024; median 1.85 vs 1.26 cm, t-test p = 0.1673, respectively). Hybrid-capture sequencing of DNA viruses (*22*) was performed on dcLN FNAs, and blood and saliva were collected for further viral assays. Clinical features of MS patients, including the Expanded Disability Status Scale (EDSS), are shown in Table 1, and further information on study subjects and assays is presented in Data file S1.

**Fig. 1.**
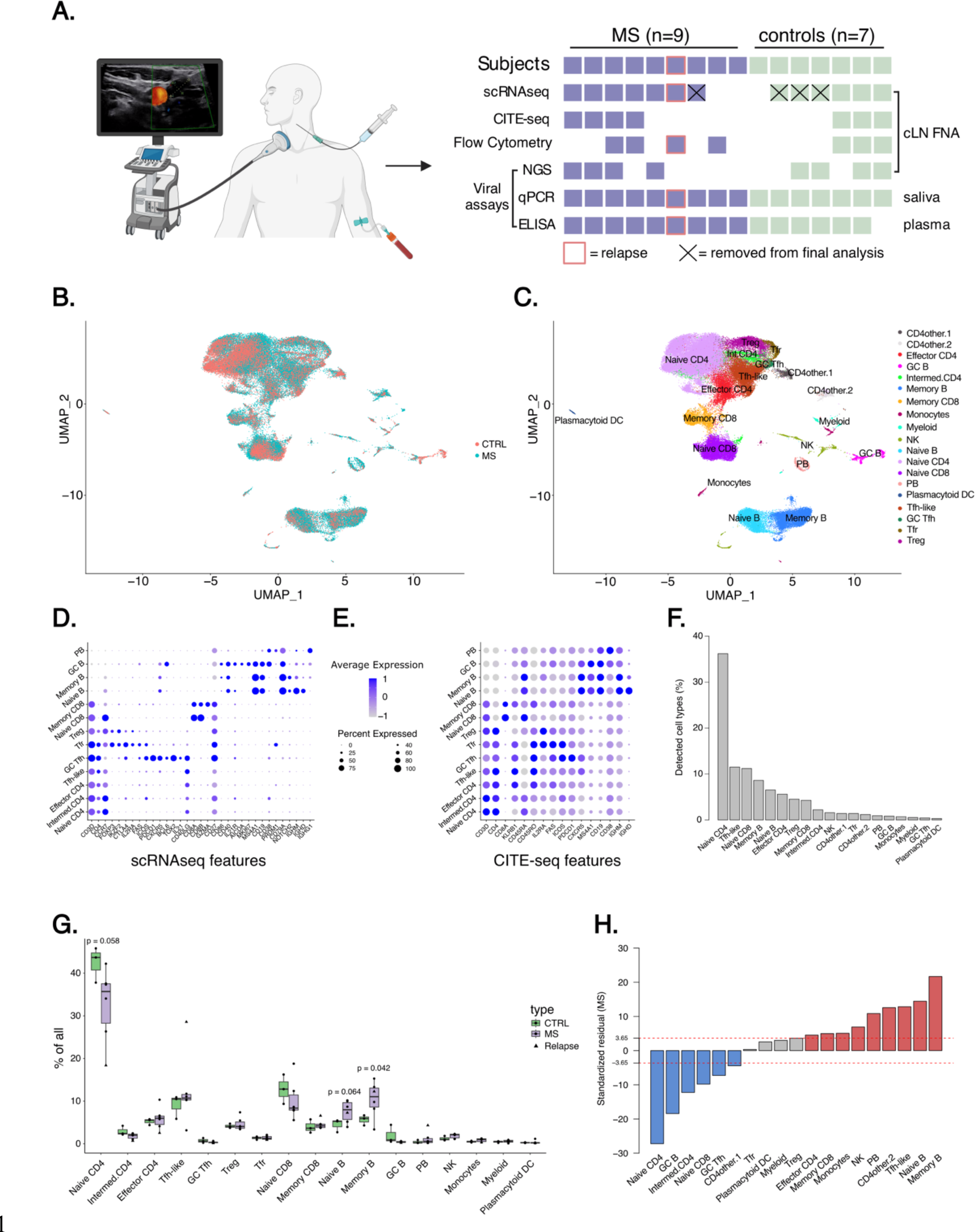
Skewed B and T cell populations in MS dcLNs highlighting enlarged memory B cell compartment **(A)** Schematic diagram of study design, where the left panel illustrates sample collection including ultrasound-guided fine-needle aspirations (FNA) of deep cervical lymph nodes (dcLN). The right panel summarizes experiments performed for each subject. (**B**) Uniform Manifold Approximation and Projection (UMAP) of identified dcLN scRNA-seq cell clusters do not show a significant batch effect. (**C**) UMAP of identified cell clusters based on a combination of automatic annotation with SingleR (the Monaco reference) and manual annotation (in-house expertise) aided by the CITE-seq data. (**D-E**) Dotplot of key scRNA-seq (D) and CITE-seq (E) marker expression profiles in identified T and B cell subsets. (**F**) Bar plot of dcLN cell type proportions. (**G**) Box plot of cell type proportions in MS patients compared to controls using an unpaired t-test (p-values < 0.1 are shown). The MS patient with an active relapse is highlighted with a triangle. (**H**) Bar plot of standardized Pearson residuals of cell subset numbers using Chi-squared test. Increased (red) and decreased (blue) cell subsets in MS patientś dcLNs are highlighted compared to healthy controls. Dashed red lines at ± 3.65 correspond to Bonferroni corrected p-values (< 0.00013) in the null distribution of standardized residuals. Abbreviations: GC, Germinal center; NK, Natural killer; PB, plasmablast; Tfh, T follicular helper, Tfr, T follicular regulator; T reg, regulatory T cell.

**Table 1:** Characteristics of MS patients.

| ID | Age | Sex | Other diseases | MS type | MRI Barkhoff | Time from onset (months) | Time from last relapse | Time from ivMP (months) | LN size (cm) | EDSS at sample collection | EDSS at 1 year follow up |
| --- | --- | --- | --- | --- | --- | --- | --- | --- | --- | --- | --- |
| MS001 | 20-29 | M | asthma, pollen allergy | RRMS | 4/4 | 8 | 8 | - | 2,06 X 0,63 | 0 | 0 |
| MS002 | 40-49 | M | no | RRMS | 4/4 | 33 | 3 | 3 | 2,00 x 0,22 | 2.0 | 1.0 |
| MS003 | 30-39 | F | no | RRMS | 3/4 | 4 | 4 | - | NA | 0 | 0 |
| MS004 | 30-40 | F | no | RRMS | 3/5 | 4 | 4 | - | NA | 1 | 1 |
| MS005 | 20-29 | F | asthma, migrane, mild ankylosing spondylitis | RRMS | 3/4 | 4 | 4 | - | 2.2 x 0.53 | 0 | 0 |
| MS006 | 40-49 | M | no | RRMS | 3/4 | 75 | 6 | 6 | NA | 1.0 | 0 |
| MS007 | 40-49 | M | no | RRMS | 4/4 | 36 | 3 | 3 | 1.04 X 0.58 | 2.5 | 5.0 |
| MS010 | 30-39 | F | no | PPMS | 4/4 | 24 | 10 | 12 | 0.88 X 0.35 | 6.0 | 6.0 |
| MS011 | 30-39 | M | no | RRMS | 4/4 | 48 | 0 | - | 1.85 X 0.58 | 3.5 | 2.0 |

Abbreviations: ivMP, intravenous methylprednisolone; EDSS, Expanded Disability Status Scale; LN, lymph node

After excluding samples with an abnormally low number of cells due to reported technical issues in 10x runs, scRNA-seq data from six MS patients and three healthy controls containing a total of 73,074 cells, with an average of 8,195 cells per MS patient and 7,968 cells for the controls, were taken for further analysis. All cells were merged for combined cluster analysis in Seurat (*23*), and the resulting clusters contained cells from each sample without evident batch effect (Fig. 1B). We compared also cell clusters between the batches (Data file S1) and observed that resulting clusters were well aligned across library preparation and sequencing batches showing no specific batch effect (see Methods and Fig. S1A-E) (*24*). We performed cell type determination using a combination of automatic annotation with SingleR (*25*) using the Monaco reference (*26*) and manual annotation aided by the CITE-seq data that we had for 4/6 MS patients and all (3/3) healthy controls (Fig. 1C). Manual cell type determination was used especially for T and B cell subsets using a combination of key markers on both transcript and protein levels (Fig. 1D-E, Data file S2-3, see Supplementary Material for details). Most of the dcLN cells were either T or B cells, whereas natural killer (NK) cell and myeloid cell subsets accounted for less than 5% of all identified cells (Fig. 1F).

Two Tfh clusters were identified and characterized by canonical *CXCR5* expression (*27*), of which the other cluster was in the minority (≈ 0.5% of all cells). Given the abundant expression of *BCL6* (*28*), *ICOS* (*29*), *TOX2* (*30*), *PDCD1* (*31*), *SH2D1A* (*SAP*) (*32*), and *IL21* (*33*), this smaller cluster likely represents a more active phenotype of Tfh (here GC Tfh) cells interacting with GC B cells (*34*) (Fig. 1D, Data file S2). Instead, the other Tfh cluster, which was the second most prevalent of all cells (≈ 11%) after naïve CD4+ T cells (≈ 40%, Fig. 1F), could reside outside the GC given the lower *BCL6* expression (*34*, *35*) and was annotated as Tfh-like cells (Fig. 1D).

### Increased proportion of memory B cells and decreased GC B cells and GC Tfh cells in dcLNs of MS patients

We noted a significantly increased proportion of memory and a marginally elevated proportion of naïve B cells in MS patients compared to healthy controls (unpaired t-test p = 0.042 and log2 fold change (log2fc) = 0.88 for memory B cells; p = 0.064, log2fc = 0.75 for naïve B cells, Fig. 1G, Data file S4). Next, we pooled all cells by subject group and compared the overall cell type frequency distribution between MS patients and healthy controls. The cell type frequencies, representing cellular compositions in dcLNs, were significantly different between MS patients and controls (Pearson’s Chi-squared test, p-value < 2.2e-16). Furthermore, evaluation of the standardized Pearson residuals suggests that in addition to memory and naïve B cells, PBs and Tfh-like cells were significantly increased while naïve and intermediate CD4s, GC B cells, naïve CD8s, and GC Tfh cells were significantly decreased in MS patients compared to healthy controls (Fig. 1H, each having extreme standardized Pearson residual values above ±3.65, corresponding to Bonferroni corrected p-values < 0.00013 in the null distribution of the standardized residuals). Tfr cells, which share transcriptional properties with Treg and Tfh cells and differentially expressed *FOXP3, IKZF2, CTLA4* on transcript level and PDCD1, and ICOS on both transcript and protein level (*19*, *36*) (Data file S2-3), did not differ between MS patients and healthy controls. Decreased GC B cells and GC Tfh cells combined with a simultaneous expansion of memory B cells suggest an imbalance of the GC reaction in MS.

We also conducted differential expression (DE) analysis for pseudobulked cell types. The highest number of DE genes (DEG) in MS samples compared to controls were observed in naïve CD4 T cells followed by plasmablasts, NK cells, and GC B cells (Fig. S1F, Data file S5). In MS patients, plasmablasts upregulated several immunoglobulin (IG) heavy and light chain genes compared to healthy controls. GC B cells upregulated *CCDN2* (*cyclin D2*, Padj = 0.024, log2fc = 1.97) and downregulated *LYN* (Padj = 0.024, log2fc = 0.93) in the MS group as compared to controls. CCDN2 is upregulated during positive selection in GCs (*37*), whereas loss of Lyn has been linked with a lupus-like autoimmune disease with hyperactive B cells (*38*). Although statistically not significant, the expression of canonical B cell marker *CD19,* known to regulate B cell proliferation in GC reaction (*39*), was decreased in GC B cells of MS patients (Padj = 0.07, log2fc = -1.12) in transcription level, and also in protein level (Padj = 5.9e-18, log2fc = -0.47) together with CR1 (Padj = 6.7e-12, log2fc = -0.47) and CR2 (Padj = 1.4e-17, log2fc = -0.55) (Fig. S1G). Interestingly, the CR1 and CR2 expression on B cells is also decreased in systemic lupus erythematosus and rheumatoid arthritis (*40*). Memory CD8+ T cells were upregulating an IFNγ modulator *DKK3* (*Dickkopf-3*, Padj = 0.0015, log2fc 2.12), *TIGIT* (Padj = 0.038, log2fc = 0.59), and cytotoxic granzymes A and K in MS patients compared to controls (GZMA, Padj = 0.038, log2fc = 0.44; GZMB, Padj = 0.0028, log2fc = 0.50). *DKK3* is upregulated in EAE (*41*) and *TIGIT* has been shown to promote CD8+ T cell exhaustion (*42*).

### Double-negative memory B cells are increased in MS patients

To further characterize the B cell compartment where we found most differences in abundance, we sub-clustered B cells. We identified similar B cell subsets as presented in Fig. 1C, in addition to a T cell cluster initially clustered with memory and naïve B cells due to a similar expression profile (Fig. S2A, Data file S6). The proportion of naïve B cells was increased in MS dcLNs after removing the remaining T cells (p = 0.035 and log2fc = 0.82, Fig. S2B, Data file S7). Then we sub-clustered the memory and naïve B cells presented in Fig. S2A and identified clusters of CD27 positive and IgD negative switched memory (SM), CD27 and IgD positive unswitched memory (USM), and CD27 and IgD low double negative (DN) B cells in addition to naïve B cells with the combination of both transcript and protein level information (Fig. 2A-B, Fig. S2C, Data file S8). MS patients had elevated proportions of the USM, SM, and naïve B cells, yet only the proportion of DN B cells was significantly larger (% of B cells, p = 0.041 and log2fc = 1.82, Fig. 2C; % of all cells, p = 0.048 and log2fc = 2.51, Fig. S2D, Data file S9). Of note, DN B cells expressed less CXCR5 than SM and USM B cells at both RNA and protein levels (Fig. 2B, Fig. S2C), indicating that DN B cells could have been derived extrafollicularly (*43*, *44*). Despite being limited by cell numbers from dcLNs, in a subset of samples with higher cell yields, we also performed flow cytometry analysis of the B cell compartment (see Methods, Fig. S2E). We adapted the gating strategy from (*45*) and observed that the proportion of GC B cells (CD19+CD20+CD38++CD10+IgD-) was reduced in MS patients compared to controls and the proportions of memory (CD19+CD20+CD27+) and naïve (CD19+CD20+IgD+CD27-CD38-) B cells were increased, however, due to the small sample size, these results did not reach statistical significance (Fig. S2F). Moreover, when dissecting the memory B cell compartment, the proportions of SM (CD19+CD20+CD27+IgD-IgM-CD38-) and DN (CD19+CD20+CD27-IgD-IgM-CD38-) B cells were higher in MS patients, however again without statistically significant difference to healthy controls (Fig S2F). The flow cytometry results align with our findings from the scRNAseq/CITEseq data, corroborating our observations of a larger memory B cell compartment together with fewer GC B cells in the dcLNs of MS patients.

**Fig. 2.**
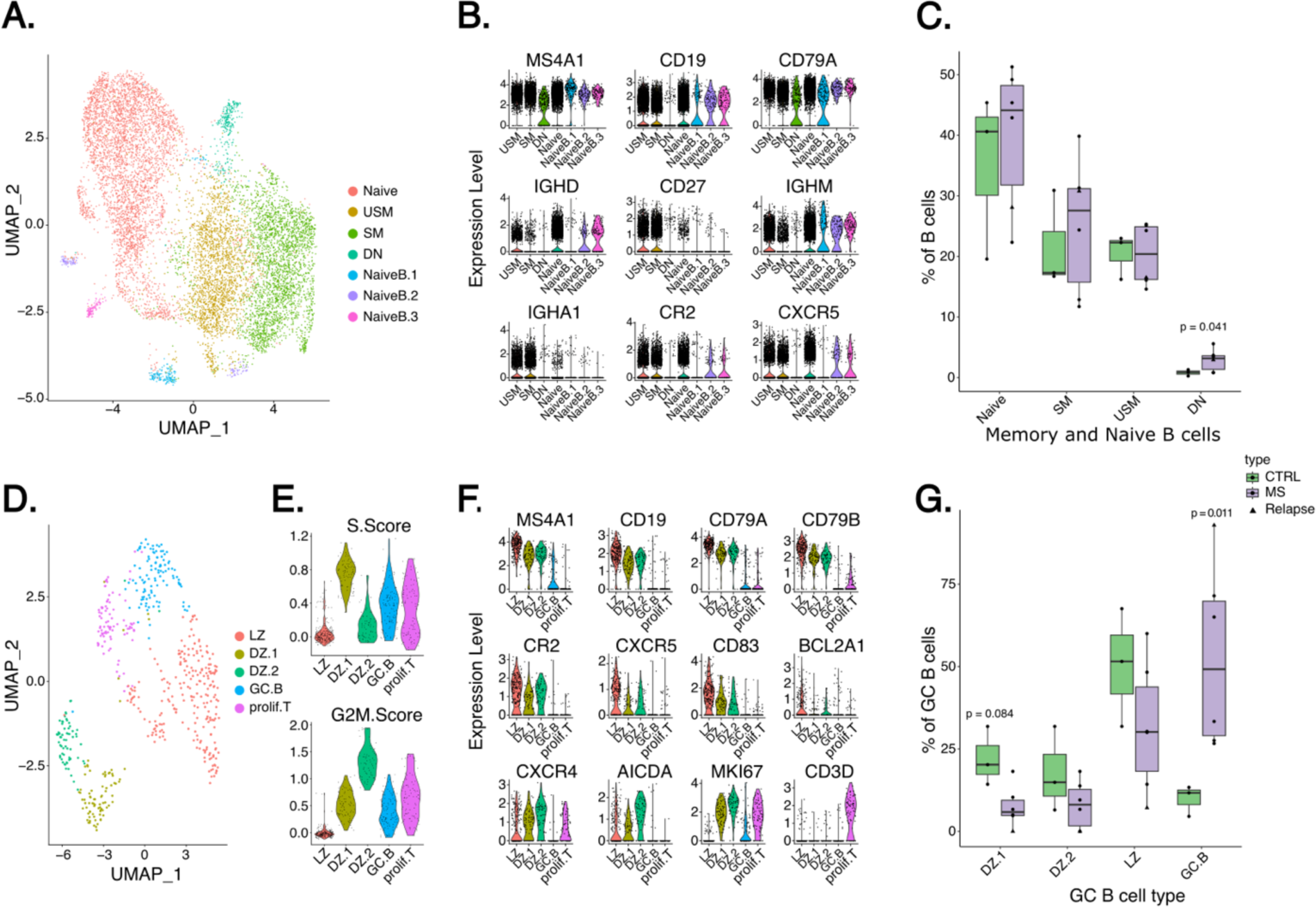
Further analysis of the B cell compartment in MS dcLNs shows increased double negative B cells (**A**) UMAP of further clustered naïve and memory B cells. (**B**) Violin plot presenting the expression of key transcripts used to identify naïve, switched memory (SM; IgD-, CD27+), unswitched memory (USM; IgD+, CD27+), and double negative (DN) B cells. (**C**) Box plot of memory and naïve B cell subsets (% of all B cells in a sample) between MS patients compared to controls. (**D**) UMAP of further clustered GC B cells. (**E**) S (upper panel) and G2M (lower panel) scores of GC B cell clusters were obtained using Seurat. (**F**) Violin plot presenting the expression of key transcripts used to identify GC B cell subsets: light zone (LZ), dark zone (DZ.1, DZ.2), and undefined GC B cell cluster (GC.B). *CD3D* expression highlights proliferative T cells (prolif.T). (**G**) Box plot of GC B cell subsets (% of all GC B cells in the sample) between MS patients and controls. In (C) and (G), an unpaired t-test was used (p-values < 0.05 are shown), and the patient with an active relapse is highlighted with a triangle.

Next, we sub-clustered the GC B cells to study the GC reaction more thoroughly. We identified *CD83* and *BCL2A1* expressing light zone (LZ) B cells (*46*, *47*), *CXCR4*, *AICDA*, and *MKI67* expressing dark zone (DZ1, DZ2) B cells (*46*, *47*), and an unidentified GC B cluster (GC.B) characterized with *MS4A1/CD20* and *MKI67* and negligible *CD3* expression (Fig. 2D-F, Data file S10). Active proliferation is a defining characteristic of GC B cells, and that is why no cell-cycle correction was performed and explains why the sub-clustering of GC B cells also contained a small cluster of proliferative T cells (Fig. 2G). Supporting the imbalanced GC reaction, we noticed that within the B cell compartment, the proportion of GC B cell subsets of all B cells showed a lower trend in MS patients (Fig. S2G). Within GC B cells, the GC.B cluster was significantly increased in MS dcLNs compared to healthy controls (p = 0.011 and log2fc = 2.42, Fig. 2G, Data file S11). As expected, pathway analysis of cluster-defining DEGs (Padj < 0.05) with GO biological processes showed a significant over-representation of pathways related to B cell activation and differentiation among upregulated genes of LZ B cells and replication among those of DZ B cells. Interestingly, the GC.B cluster, which was increased in MS patients, showed an over-representation of ribosomal genes related to viral transcription and viral gene expression (Data file S1).

### Cellular interactions between follicular T cells and GC B cells

Since MS patients had an increased proportion of naïve B cells, memory B cells, and PBs, while GC B cell and GC Tfh proportions were decreased compared to healthy controls, we hypothesized that the interaction between T and B cell subsets is altered. To study intercellular interactions, we used NicheNet (*48*), which links transcripts of ligands in sender cells to their transcript targets in the receiver cell. Principally developing GC B cells rely on signals from Tfh via CD40 – CD40L interaction to undergo SHM and class-switching (*49*). Similarly, the most active CD40 – CD40LG interactions for GC B cells were predicted between GC Tfh and Tfh-like cells (Fig. 3A). Integrins prolong interactions between T and B cells (*50*), and interestingly *CD40LG – ITGB1* (integrin beta 1) interaction was observed only between GC Tfh and GC B cells emphasizing their crosstalk in GCs. GC Tfh cells were the only CD4+ T cells which interacted with GC B cells via *CD28 – CD86* and *CD28 – CD80*, of which the latter is reported to be crucial for T-B interactions in GC responses (*51*). Contrary to our hypothesis, the above interactions between GC B cells and Tfh cells were similar for MS patients and controls when compared using MultiNicheNet (*52*) (Fig. S3A). We observed a slight increase in *galactin-1 (LGALS1) – lectin receptor CD69* interaction between GC B and Tfh cells in MS patients (Fig. S3A), of which galactin-1 is shown to favour PB differentiation (*53*). We did not observe *IL21 – IL21R* interaction between GC B cells and GC Tfh cells, however, this could have been affected by the small proportions of both cell subsets. Instead, *IL21 – IL21R* interaction was increased between GC Tfh and naïve B cells, suggesting that part of the naïve B cells could have already been dedicated for the GC reaction (*33*) (Fig. 3A). In line with this, *CD40LG – CD40* interaction between GC Tfh/Tfh-like cells and naïve B cells was decreased in MS patients compared to GC B cells (Fig. S3A). The naïve B cells dedicated to the GC reaction could potentially receive less necessary proliferative signals from Tfh cells in MS patients, which could lead to an altered GC response.

**Fig. 3.**
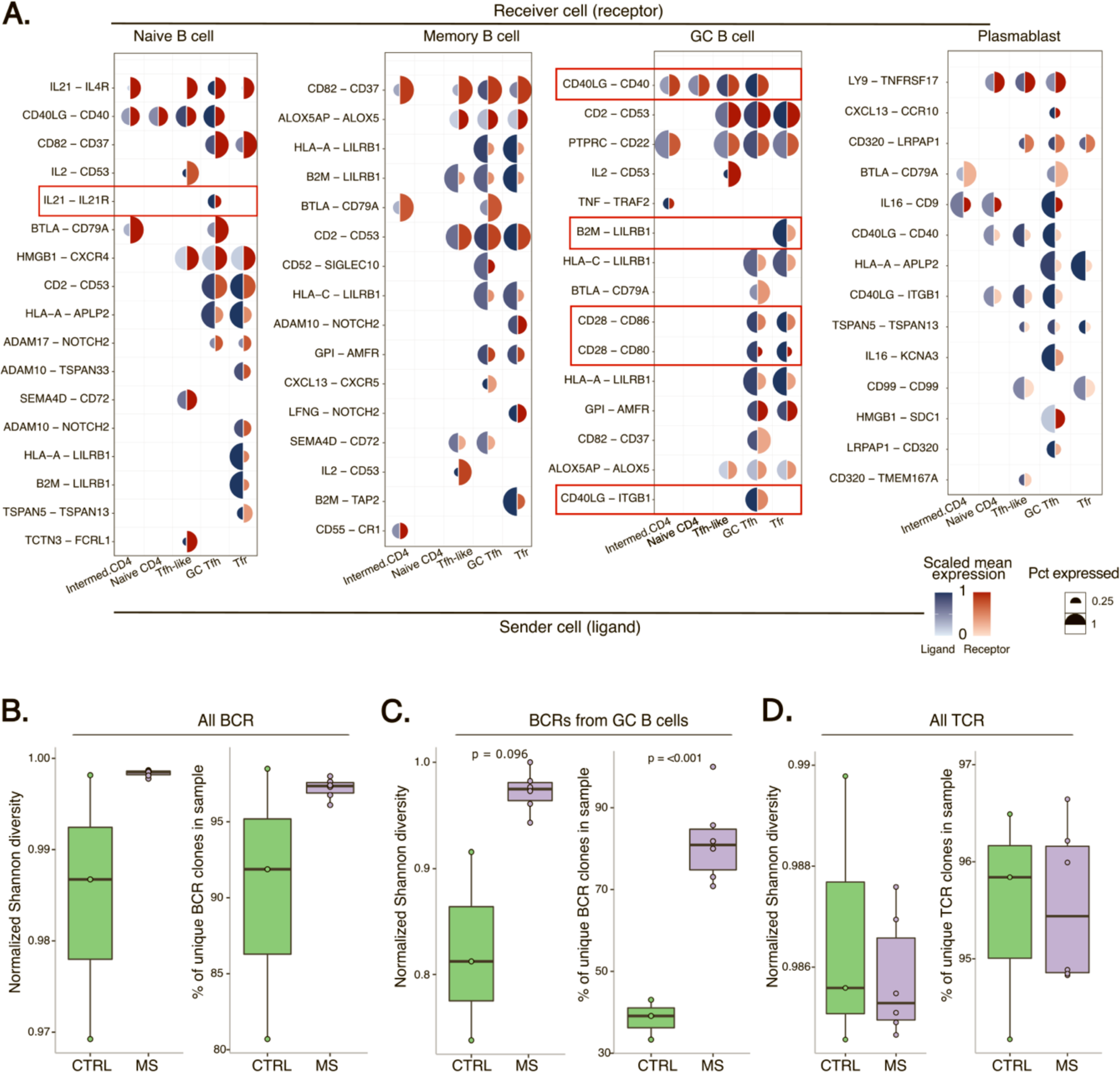
GC BCRs in MS dcLNs are less clonal **(A)** Mushroom plot showing intercellular communication of helper T and Tfr cells (ligand) between naïve, memory GC B cells and plasmablasts (receptor) using NicheNet. The colour of red and blue corresponds to the scaled mean (from 0-1) expression of ligand/receptor, while the size of the circle corresponds to a percentage of cells expressing a particular transcript. Red rectangles highlight intercellular communications emphasized in the Results. Diversity of (**B**) all BCRs, (**C**) BCRs of GC B cells, and (**D**) all TCRs presented by boxplots from two different metrics of measuring repertoire diversity: normalized Shannon entropy (0 corresponds to monoclonal BCRs/TCRs, and 1 corresponds to all BCRs/TCRs would be diverse) and percent of unique clonotypes. In each case, an unpaired t-test was used to compare diversity estimates between MS patients and controls (p-values < 0.1 are shown with p-values < 0.05 and < 0.1 considered significant and marginally significant respectively).

The precise mechanisms of how Tfr cells control the GC reaction and selection of Tfh and GC B cell clones are largely unknown. Here, Tfr cells were predicted to interact with GC B cells via *CD28 – CD80/CD86* (Fig. 3A), which is needed for Tfr differentiation (*54*). We did not observe CTLA4-mediated inhibition of CD80/CD86 costimulatory signals between Tfr and GC B cells, which has been thought to play a key role in the regulation of GC reaction (*55*). B cell proliferation, memory responses, and CXCR5 expression of GC B cells are downregulated via HLA-G – LILRB1 (also known as ILT2) interaction between T and B cells (*56*). B2M (beta-2-microglobulin) constitutes the light chain of HLA-G, and *B2M – LILRB1* interaction was predicted only between Tfr and GC B cells (Fig. 3A). Possibly B2M – LILRB1 interaction is one mechanism for Tfr cells to regulate GC B cell development and thus, GC reaction.

### GC B cells are less clonal in dcLNs of MS patients

We evaluated the BCR repertoire in dcLNs of MS patients and healthy controls. On average, BCRs from 1400 cells per MS patient and 882 per healthy control sample were obtained. We observed no difference in the diversity of the total BCR repertoire (Fig. 3B), nor in the BCR diversity of naïve B cells, memory B cells, or plasmablasts between patients and controls (Fig. S3B). However, the BCR diversity of GC B cells was significantly higher in MS patients when compared to healthy controls (Fig. 3C). To confirm this, we used the Lanz et al. (*8*) definition for BCR clonotypes (see Methods) where we still observed significantly increased diversity only in the GC B cell compartment of MS patients (Fig. S3C). In summary, while we could detect a lower proportion of GC B cells in MS dcLNs (Fig. 1H), their standardized BCR diversity was higher than in healthy controls.

### TCR repertoire in dcLNs is similar between MS patients and controls

From T cells we obtained single-cell paired TCRαβ data for an average of 6124 cells per MS patient and 6702 for healthy control, corresponding to 5851 and 6384 average unique TCR clonotypes, respectively (Fig. S3D). Comparison of TCR diversity in the entire dcLN-paired-TCRαβ repertoire, with and without downsampling to the same read- depth, showed no statistical difference in TCR clonality between MS and healthy controls (Fig. 3D). We also found no statistical difference in repertoire diversity within specific T cell subpopulations between patients and controls (Fig. S3E). To determine the possibility of more TCR sharing among MS patients, we also compared TCR amino acid level overlap rates for every possible pair between patients and controls, however, no evidence for increased TCR overlap in MS dcLNs was found (Fig. S3F).

Given the decreased GC B cell clonality and proportion in MS dcLNs, we examined follicular T cells closer. Only a few TCRs of Tfh-like, GC Tfh, and Tfr cells were shared with TCRs from other T cell subsets within each individual, with no difference between MS patients and controls (Fig. S3G). Tfr cells shared TCR clones with both Treg (in 3/6 MS patients and 1/3 controls) and Tfh-like cells (in 1/6 MS patients and 1/3 controls), indicating that Tfr cells had developed from both cell types. A trajectory analysis of the scRNA-seq data from Tfh-like, GC Tfh, Tfr, and Treg cells using slingshot (*57*) showed a single developmental trajectory encompassing from Tfh-like cells via GC Tfh and Tfr to Treg cells, suggesting that Tfr cells had developed from both Tfh and Treg cells, in line with recent findings (Fig. S3H) (*58*).

To compare the antigen-driven TCR repertoire in the patients’ dcLNs to controls, we scanned the VDJdb database for possible specificity of all our TCRαβ amino acid clonotypes using tcrdist3 (see Methods). Predicted T-cell-specificity was higher in MS patients towards epitopes of M.tuberculosis in Naïve CD4s (p-value=0.03, log2fc = 0.5), Influenza A (p-value = 0.003, log2fc = 3) and EBV (marginally with p-value = 0.08 & log2fc = 0.56) in GC Tfh cells (Fig. S3I), however, these were insignificant after we filtered for HLA restriction of target TCRs (retaining HLA class 2 restricted hits for CD4+ T cells, and HLA class 1 restricted hits for CD8+ T cells).

### Higher coverages of EBV, HHV-6B and HHV-7 in dcLNs and viral loads of EBV in the saliva of MS patients

All MS patients were seropositive for EBV viral capsid antigen (VCA) IgG (9/9), whereas only five of the six controls were EBV VCA IgG seropositive. Of the EBV VCA IgG seropositive individuals, MS patients had higher EBV VCA IgG titer than healthy controls (p = 0.019, Fig. S4A). None of the subjects were seropositive for EBV VCA IgM.

Next, by using hybrid-capture sequencing, we searched for common human DNA viruses from fine needle aspirations of dcLNs (nMS=4, nhc=4, Fig. 1A). The prevalence of EBV (75% vs 50%, human-herpesvirus-6B (HHV-6B, 50% vs 0%), and HHV-7 (100% vs 75%), but also the total number of unique viral reads covering the reference sequences, which correlates to the viral copy numbers (*59*), was higher in MS patients compared to controls (Fig. 4A, Table 2). The coverages of HHV-7 were particularly higher in MS patients, from whom we were able to reconstruct two near-complete viral genomes. Three out of four MS patients were positive for parvovirus B19 (B19V) compared to 100% of controls. Equal prevalence (50%) of Merkel cell polyomavirus was detected in both MS patients and healthy controls. Importantly, MS patients and healthy controls clustered into separate clusters based on the breadth coverage profiles of the above-tested viruses (Fig. 4A). We aligned the scRNA-seq data to reference viral genomes available at ViruSITE (*60*) and NCBI (*61*) databases (see Methods) including EBV genome to identify cells producing viral transcripts (*62*). However, we did not find any cells producing transcripts of EBV, or other herpesviruses.

**Fig. 4.**
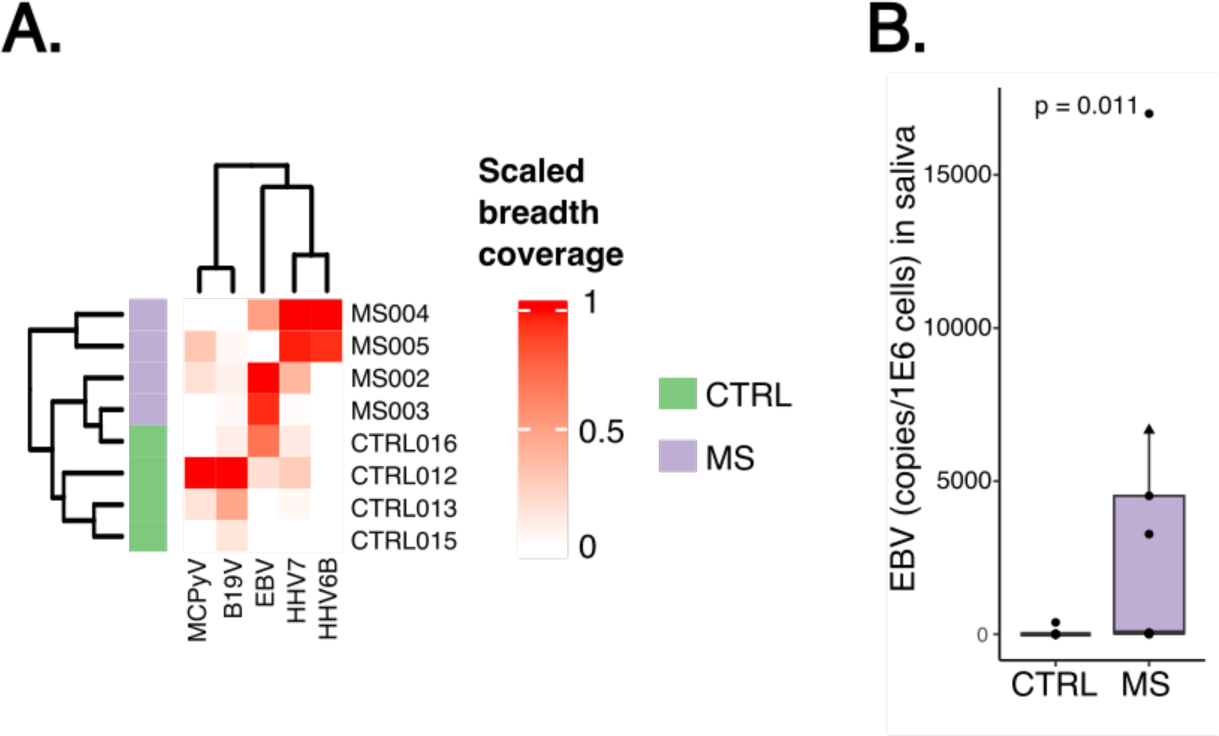
EBV, HHV-6B and HHV-7 show higher breadth coverage in dcLNs and EBV viral load is increased in MS patients compared to controls (**A**) Heatmap illustrates the presence and the percentage of viral reads covering the EBV, HHV-6B, HHV-7, B19V, and MCPyV reference sequences (breadth coverage) in dcLNs determined using hybrid-capture sequencing. Viral breadth coverages were normalized from 0 to 1 separately for each virus, followed by agglomerative hierarchical clustering using Euclidean distance and Ward.D2 linkage. Dendrograms show the clustering of samples based on the breadth coverage profiles of the above-tested viruses. Negative samples for tested viruses are presented in white. (**B**) Box plot of EBV copies per 1 million cells in the saliva of MS patients compared to controls. EBV copy numbers were determined using qPCR. An unpaired t-test was used, and the patient with an active relapse is highlighted with a triangle.

**Table 2:** Results from the viral assays.

|  |  | Plasma | Saliva (copies / 1*10^6 cells) |  |  | cLN (breadth of coverage in %) |  |  |  |  |
| --- | --- | --- | --- | --- | --- | --- | --- | --- | --- | --- |
| ID | type | EBV VCA IgG (RU/ml) | EBV | HHV6B | HHV7 | EBV | HHV-6B | HHV-7 | MCPyV | B19V |
| CTRL008 | CTRL | 0 | 0 | 82 | 32400 |  |  |  |  |  |
| CTRL009 | CTRL | 104,59 | 387 | 132 | 77,1 |  |  |  |  |  |
| CTRL012 | CTRL | 151 | 0 | 0 | 347 | 5,13 | 0 | 26,4 | 22.4 | 89,5 |
| CTRL013 | CTRL | 63,5 | 0 | 17,9 | 2210 | 0 | 0 | 3,8 | 3.4 | 41,2 |
| CTRL014 | CTRL | 137 | 0 | 38,2 | 525 |  |  |  |  |  |
| CTRL015 | CTRL | 146,3 | 0 | 334 | 25300 | 0 | 0 | 0 | 0 | 12,3 |
| CTRL016 | CTRL |  | 0 | 491 | 211000 | 20,8 | 0 | 11,4 | 0 | 8,4 |
| MS001 | MS | 172,86 | 3270 | 33,8 | 2030 |  |  |  |  |  |
| MS002 | MS | 136,26 | 52,8 | 314 | 375 | 29,9 | 0 | 36,8 | 3,5 | 6,2 |
| MS003 | MS | 179,04 | 0 | 69,2 | 11800 | 28,2 | 0 | 1,84 | 0 | 3,1 |
| MS004 | MS | 223,73 | 26,2 | 297 | 3680 | 14,7 | 10,12 | 98,6 | 0 | 0 |
| MS005 | MS | 194,59 | 17000 | 219 | 74100 | 0 | 9,5 | 95,67 | 6,7 | 3,5 |
| MS006 | MS | 179,72 | 4520 | 0 | 221 |  |  |  |  |  |
| MS007 | MS | 209,72 | 10,4 | 15,2 | 2350 |  |  |  |  |  |
| MS010 | MS | 153,48 | 37 | 0 | 1890 |  |  |  |  |  |
| MS011 | MS | 177,19 | 6660 | 389 | 154000 |  |  |  |  |  |

Abbreviations: EBV, Epstein-Barr Virus; HHV, human herpes virus; MCPyV, Merkel cell polyomavirus; B19V, parvovirus B19

Using multiplex qPCR HERQ-9 quantification we searched for EBV DNA from the saliva as a sign of EBV reactivation. We found three out of nine HHVs in the saliva of MS patients and controls: EBV, HHV-6B, and HHV-7. Herpes simplex virus-1 (HSV-1), HSV-2, varicella-zoster virus, CMV, HHV-6A or Kaposís sarcoma-associated herpesvirus were not detected in any of the samples. EBV was found from saliva in all except one of the MS patients in contrast to only one of the controls, and the viral loads in MS patients were significantly higher (p = 0.011) (Fig. 4B). Surprisingly, the MS patient negative for EBV DNA in dcLN had the highest EBV viral load in saliva. HHV-6B was found in most samples and HHV-7 in all without significant differences in the viral loads for either of these viruses between groups (Fig. S4B).

### Patient with an active MS relapse exhibits EBV targeting memory CD8 cells, skewed follicular T cell ratios, and negligible GC B cells

Immunological processes during active MS relapse are poorly understood, partly because treatment is usually started soon after hospital admission. We managed to obtain dcLN FNA from one patient during hospitalisation for an active MS relapse before immunomodulatory treatments were started. The patient (MS011) had no other known diseases besides MS nor an immediate history of infections and exhibited weakness of the right leg and diplopia with EDSS of 3.5 in the clinical evaluation during the relapse (Table 1). The patient’s plasma EBV VCA IgG concentration and copy numbers of HHVs in saliva were comparable to MS patients (Table 2).

When comparing cell abundancies from dcLNs, we observed that the patient had the highest proportions of PBs, Tfh-like, and memory CD8 T cells, and the lowest proportions of naïve, intermediate, and effector CD4, GC Tfh, Tfr, and GC B cells across all subjects (Fig. 1G, Fig. S5A). The abundance of memory and naïve B cells were similar to those in other patients. The ratio of GC Tfh to Tfh-like cells was reduced, while the total Tfh to Tfr ratio was significantly increased (Fig. 5A), both likely due to the drastic increase of Tfh-like cells (Fig. S5A). Moreover, the patient had the lowest proportion of DZ and LZ B cells of all B cells. Within the GC B cell compartment the cluster of GC.B cells, characterized by upregulation of transcripts involved in pathways of viral transcription and viral gene expression, was significantly increased (Fig. 2G, S2D, S5B).

**Fig. 5.**
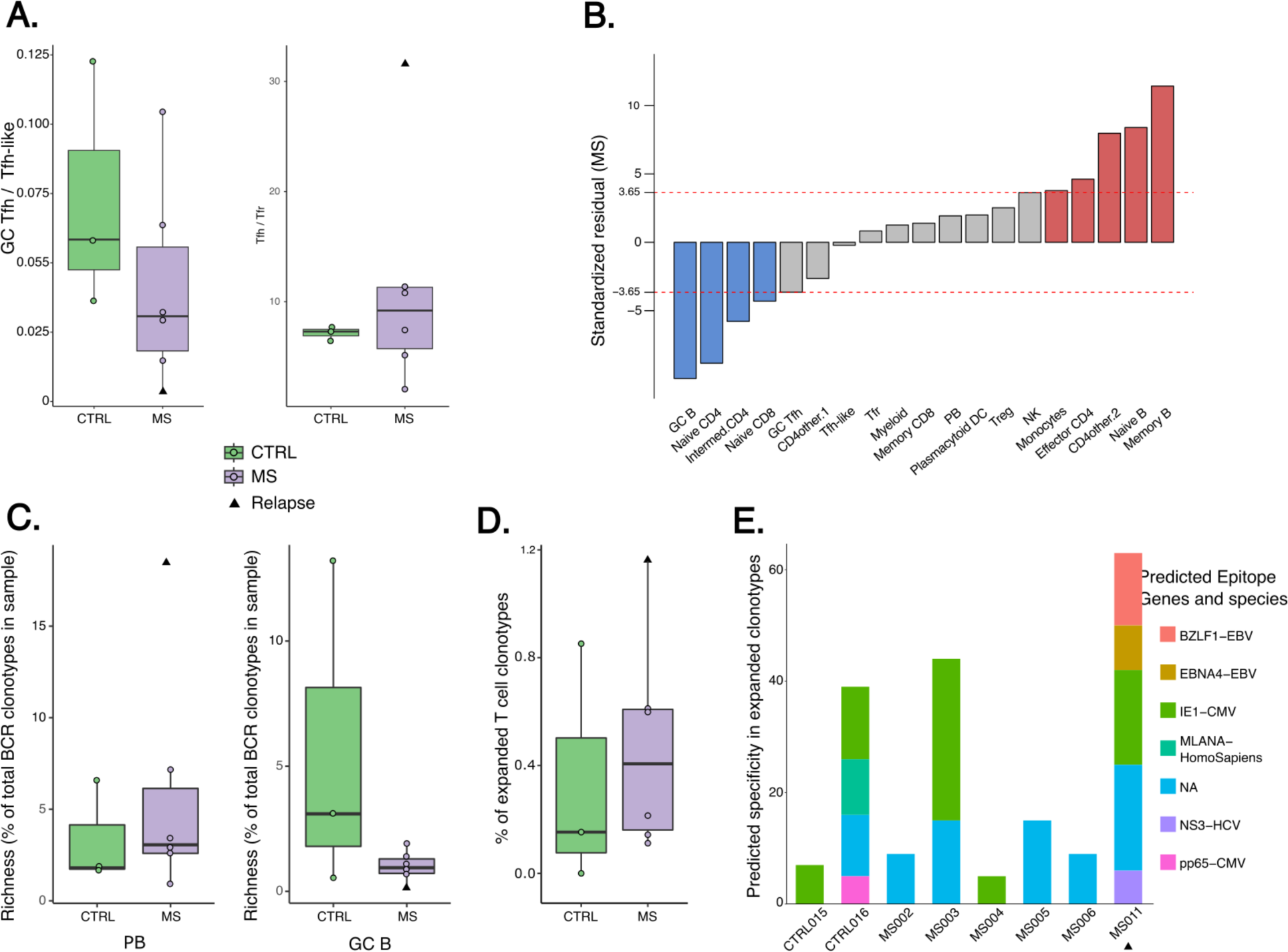
MS patient with an active relapse exhibits altered follicular cell ratios and clonally expanded EBV targeting CD8+ T cells **(A)** Box plots of GC Tfh / Tfh-like (left) and Tfh / Tfr ratios (right) in the MS patients compared to healthy controls. Tfh is a sum of Tfh-like and GC Tfh cell counts. **(B)** Bar plot of standardized Pearson residuals of cell subset numbers between controls and MS patients without the patient with a relapse using Chi-squared test. Increased (red) and decreased (blue) cell subsets in MS patients compared to healthy controls are highlighted. Dashed red lines at ± 3.65 correspond to Bonferroni corrected p-values (< 0.00013) in the null distribution of standardized residuals. **(C)** Box plots of plasmablast (left) and GC B cell (right) BCR richness (BCRs of total BCR clonotypes in each sample). (**D**) Box plot of expanded T cell clonotypes of all T cell clones (%) in each sample. (**E**) Stacked bar plot of predicted specificity in expanded T cell clonotypes. The VDJdb database was scanned for TCRab and b clonotypes using tcrdist3. In A and C-E, the patient with an active relapse is highlighted with a triangle.

We also redid analysis on cellar abundancies between patients and controls after excluding the patient with acute exacerbation to evaluate how much of the results were affected by the acute case. After excluding the acute relapse patient, non-treated MS patients still had significantly increased memory and naïve B, and effector CD4 T cells, decreased GC B, naïve CD4, and CD8, and intermediate CD4 T cells, and marginally decreased GC Tfh cells (Fig. 5B). To conclude, the alterations of lymphocytes characterized with larger memory B cell compartment and smaller proportion of GC B cells in dcLNs seem more pronounced in early MS during a relapse.

The patient had the highest number of all BCRs and PB BCRs (Fig. 5C), whereas the lowest of GC B cell BCRs (5 GC B cell BCRs of 1758 detected BCRs, detected BCRs in all samples ranged from 599 to 1758). On the TCR side, while the relative frequency of expanded T-cell clonotypes was not statistically different between the patient groups, the largest percent of cells from expanded TCR clonotypes was seen in the patient with relapse (1.16% of all T cell clonotypes were expanded, the median in MS patients is 0.40% and in controls 0.15%; paired TCR clonotypes with frequencies of over 0.001 of all clonotypes within each sample were considered expanded), of which the most (84%) were memory CD8 T cells (Fig. 5D). Notably, the majority of the clonally expanded T cells from the patient were predicted to be EBV specific, whereas no EBV specific TCRs were seen in T cells of other subjects (Fig. 5E). We validated and refined the TCR predictions with an orthogonal method TCRGP, which has outperformed the original tcrdist algorithm (*63*). With TCRGP, we confirmed the enrichment of clonally expanded CD8+ TCR specific to EBV antigen EBNA3B in the patient with an active relapse (Fig. S5C, Padj = 0.0002, odds ratio (OR) = 18.80, Benjamini-Hochberg corrected one-sided Fisher’s exact test).

## DISCUSSION

Therapeutic advances during the last decades have highlighted the role of B cells and T-B crosstalk in MS pathogenesis. Examining B cell subsets at the site where B cell-mediated immunity evolves is essential for gaining a deeper understanding of the origin of MS and for elucidating why systemic B cell-depletion effectively treats the disease.

Here, to the best of our knowledge, we have provided the first-ever characterization of the single-cell immune landscape in the dcLNs of newly diagnosed MS patients by coupling scRNA-seq with CITE-seq. The B cell compartment of dcLNs is altered in MS patients with an increase in memory B cells and decreased GC B cells with reduced clonality suggesting a disturbance of GC reactions. Hybrid-capture sequencing of eukaryotic DNA virome detected EBV, HHV-6B and HHV-7 DNA more readily in the patients’ dcLNs than the controls. Moreover, using multiplex qPCR from the saliva against common herpesviruses, EBV DNA was witnessed primarily in the patients and not the controls. In addition, the MS patient sampled during an acute relapse presented with starker findings in the same B cell subsets together with clonally expanded memory CD8+ T cells whose TCRs were likely targeting EBV.

Expansion of memory B cells and plasma cells in the CSF of MS patients is well demonstrated (*8*, *16*, *64*). Simultaneously, Tfh cells are more prevalent both in CSF of MS patients and in MS animal models, where Tfh cells promote B cell accumulation and disease progression (*64*). Therapeutic effect of systemic B cell depletion is mirrored in CSF by a decrease in memory B cells, which further underlines the role of B cell dysregulation in MS. Based on studies with twins (*65*) or with paired CSF and blood samples (*64*, *66*) the B cell expansion is restricted to the CNS, whereas peripheral blood reflects the B cell compartment inadequately. Even though the expanded memory B cells are shown to derive from the dcLNs (*16*), the role of dcLNs in MS has been largely unstudied. To enlighten this, we investigated the dcLNs in MS patients and controls and discovered that the proportion of memory B cells and to a smaller degree PBs was augmented in dcLNs of MS patients. This suggests that the reported expansion of intrathecal B cells, plasma cells (*64*) and circulatory memory B cells in early MS (*67*) derive from alterations of B cell maturation in the dcLNs. In addition, MS patients had fewer and less clonal GC B cells further suggesting dysregulation in GC reaction. Also, the Tfh cells likely representing GC-resident Tfh cells (*34*) interacting with GC B cells due to expression of *BCL6* (*28*), *ICOS* (*29*), *TOX2* (*30*), *PDCD1* (*31*), *SH2D1A* (*SAP*) (*32*) were decreased in MS patients. Tentatively, GC B cells could receive less survival signals in the light zone from GC-resident Tfh cells leading to favoring the B cell differentiation toward memory B cell fate over PB fate (*49*, *68*) and to GC contraction (*69*) in dcLNs of MS patients. This would subsequently explain the decrease of GC B cells in MS patients. Interestingly, autoimmunity-linked DN (*45*) but also switched-memory B cells were increased in MS patients. Both of them could develop in extrafollicular responses of the lymph node, not only in GCs (*43*). On the other hand, Tfh cell-proportions were skewed especially in the patient with a relapse, in which Tfh-like cells, that probably remain outside the GC (GC-nonresident Tfh), were expanded. Chronic viral infections seem to favour Tfh differentiation, yet result in dysregulated Tfh – GC B cell interactions (*79*). Hence, local activation of latent EBV could be a driver for the GC-nonresident Tfh expansion during the disease relapses leading to a larger memory B cell compartment in MS patients as observed here.

Further studies on larger datasets with paired CSF samples are warranted to demonstrate whether altered GC reaction in dcLNs is the origin of clonally expanded intrathecal B cells in MS, potentially opening the door to new therapeutic strategies.

Epidemiological evidence for EBV infection as a prerequisite for MS onset is strong. However, the exact pathogenic mechanisms leading to MS after this ubiquitous viral infection are still unresolved. Molecular mimicry between CNS structures and EBV has been widely acknowledged (*8*), but why such reactions lead to CNS autoimmunity in only a minority of humans infected with EBV remains still unresolved. Findings of EBV from postmortem samples have been contradicting (*70–72*), thus an uncontrolled viral activation or replication as the cause of MS is still debatable. Interestingly, spontaneous proliferation of CD4+ T cells initiated by memory B cells has been reported in MS patients with HLA-DR15 haplotype (*73*) and is stimulated by EBV peptides (*74*).

Considering the observed expansion of memory B cells in MS dcLNs, EBV-infected B cells could be the source of uncontrolled proliferation of CD4+ T cells.

Contrary to previous results by Gieß et al (*75*), we observed EBV DNA in the saliva of adult early MS patients, possibly due to a more sensitive method. Similarly, oral shedding of EBV as a possible sign of EBV lytic cycle reactivation has been reported in pediatric MS patients (*76*). We detected EBV DNA in higher prevalence in MS patients dcLNs than in controls, however, we could not find cells producing EBV nor other aforementioned viruses by aligning scRNA-seq reads to viral genomes. Yet, it should be noted that the copies of the viruses detected by hybrid-capture sequencing were likely to be few, and the low efficiency of scRNA-seq library preparation (*77*) limits the likelihood of detecting cells with viral transcripts from the scRNA-seq data. Our findings of EBV activation lead us to speculate whether, in early MS, relapses could rise from local activation of latent EBV in dcLNs, which would not be detected systematically with serological assays (*78*). Recently, EBV-specific memory CD8+ T cells were observed in CSF of both healthy controls and MS patients, however, patientś had also central memory CD8+ T cells associated with broader EBV-specific repertoires (*80*). In our study, the patient with an ongoing relapse had expanded EBV targeting memory CD8+ T cells, and it is possible that they expand and migrate to CNS during a relapse, since this was not observed in patients without active relapses.

The understanding of GC reaction is largely based on experiments with mice, and studies with human tissues, especially in autoimmunity are few since patient recruitment and sample collection are challenging. Our dcLN data set from recently diagnosed MS patients without immunosuppressive medication is unique, however, it is restricted with the small sample size. Also, our scRNAseq experiments were limited to 10,000 cells per sample, which impeded the T and B cell clonotype analysis. The majority of the VDJdb database is based on CD8+ T cells, which could explain why we could not detect CD4+ T cell subsets targeting several viruses, such as EBV. Despite the limited cell number per sample, we studied paired TCRαβ data from an average of 6124 T cells per MS patient and 6702 per healthy control, which increases the specificity of our clonotype results. Yet, we did not find any differences in repertoire diversity within specific T-cell subpopulations when comparing MS patients to healthy controls, which suggests that clonal expansion of specific T cell clones unlikely dominates in dcLNs outside the MS relapses. The only EBV-targeting T cells were clonally expanded memory CD8+ T cells found only in the MS patient with a relapse. Using TCRGP, we confirmed that the memory CD8+ TCRs in the patient with relapse were specific to latency-associated EBV antigen EBNA3B (*7*) and increased 18.8 times compared to healthy controls.

Our results encourage the use of FNA and scRNAseq to study lymphoid tissues, including ectopic GCs in autoimmune infiltrates, in various diseases to better understand autoimmune mechanisms and development of memory B cells and plasmablasts. By investigating the single-cell immune microenvironment of dcLNs at the diagnosis of MS, we observe increased memory B cell compartment along with decreased GC B cells in comparison to healthy controls, suggesting malfunction of the GC reaction in the patients. Paralleled with findings related to EBV and the known influence of EBV on GC dynamics (*81*), our results support B cell dysregulation as a key mechanism in MS pathogenesis.

## MATERIALS AND METHODS

### Study design

We studied the cellular compartment of dcLNs in MS patients and healthy controls using ultrasound-guided FNAs. Viable single cells were isolated from the FNAs and subsequently processed for 5’ single-cell RNA sequencing and hybrid-capture sequencing of DNA viruses (*22*). Also, blood and saliva were collected from the participants for additional viral assays.

This study was approved by the Helsinki University Hospital ethical committee (decision number 5774/2020; research permit number 2363/2020). All subjects participated voluntarily and gave written informed consent according to the principles of the Declaration of Helsinki. Nine newly diagnosed MS patients and 7 age and sex-matched healthy controls were selected for this study between April 2021 and March 2022. MS diagnosis was confirmed by an experienced neurologist at the Helsinki University Hospital using the 2017 McDonald diagnostic criteria (*82*, *83*). The full diagnostic workup included evaluation by a neuroimmunologist, CSF analysis, and a head MRI. All MS patients had oligoclonal bands in CSF at the time of diagnosis. We only chose MS patients without any immunomodulatory medications. In addition, an absence of intravenous glucocorticoid infusion within 3 months before sample collection was a part of the inclusion criteria. Further patient characteristics are shown in Data file S1.

### Sampling of blood and saliva

Blood and saliva were taken 1 to 3 hours before the FNA sample. Blood samples were collected into Li-heparin Vacutainer tubes (BD Biosciences), and plasma was separated by centrifugation and archived at -80 C° until enzyme-linked immunosorbent assay (ELISA). Subjects had a minimum of a 30-minute break from any liquids or solid foods before saliva samples were obtained. The samples were collected at noon to normalize the diurnal variation of protein secretion and stored at -20 C°. EBV serology status was evaluated using Anti-EBV-CA ELISA IgG and IgM kits (Euroimmun, CAT#: El 2791- 9601 G, EI 2791-9601 M) according to the manufactureŕs instructions.

### Fine needle aspirations

Ultrasound-guided FNAs of dcLNs and subsequent processing for single-cell sequencing were performed as described previously by Turner et al (*84*). Briefly, radiologists (G.K, R.L) first measured dcLNs (Table 1), after which they performed FNAs under visualization with a 25-gauge needle. For each sample, 2 aspirations were taken and collected into 3ml of 10% FBS in RPMI 1640 followed by three 0.5 ml rinses. After one wash with 2% FBS and 0.5mM EDTA in PBS, red blood cells were eliminated using 1ml of ACK lysis (Lonza) followed by another wash using PBS supplemented with 2% FBS and 0.5mM EDTA. Single-cell RNA sequencing was performed in concentrations of 1x10^7 cells per ml in calcium- and magnesium-free PBS with 0.04% BSA solely or in parallel with the cellular indexing of transcriptomes and epitopes by sequencing (CITE-seq). CITE-seq staining was performed for a minimum of 500,000 cells using TotalSeq™-C Human Universal Cocktail (BioLegend, cat 399905) according to the manual. For the samples in which solely scRNAseq was performed, 10,000 cells were targeted for sequencing. For part of the samples (Data file S1), a flow cytometry assay was also performed to characterize B cell subsets (described further). The remaining cells were frozen in RNA Later (Thermo Fisher) according to the manufactureŕs instructions at -80 C°.

### Single-cell sequencing

scRNAseq was performed for 7/9 MS and 5/7 healthy control dcLN FNA samples of which CITE-seq was performed for 5 MS and all healthy control samples. FIMM Single-Cell Analytics core center at the University of Helsinki performed 5’RNAseq run and library preparation for scRNAseq, TCRαβ, and BCR libraries using the Chromium Single Cell 5’ (v2 Chemistry Dual Index) with Feature Barcoding technology for Cell Surface Protein and Immune Receptor Mapping (CITE-seq) as instructed by the manufacturer (10X Genomics).

The sequencing was started immediately after FNA sample preparation and libraries were sequenced on Illumina NovaSeq 6000 system (read lengths 26bp (Read 1), 10bp (i7 Index), 10bp (i5 Index), and 90bp (Read 2). The raw scRNAseq data were processed using Cell Ranger (v 2.2.0) against the human genome GRCh38. TCRαβ and BCR data were processed with Cell Ranger vdj pipeline to generate single cell V(D)J sequences and paired clonotype calling according to the manufactureŕs instructions.

### Single-cell RNA, CITE-seq, and scTCR & BCR data analysis

Analysis of scRNAseq data was performed using Seurat (v4.1.1) in R (*23*). Initial QC was performed separately for each sample in which cells in the top 2 percentile for mitochondrial gene expression and in the top 1 percentile for the number of genes detected were removed. Also, one MS sample and three healthy control samples with an abnormally low number of cells due to technical issues in 10x runs were removed. The remaining samples were combined for cluster analysis with the merge function in Seurat. Uniform manifold approximation and projection (UMAP) was then performed in Seurat using the top 50 principal components of the merged data. Graph-based clustering and cluster labelling were performed with a resolution of 1 of the FindClusters function. The resulting clusters were defined by cell types and were well mixed from all possible batches (library and sequencing batches, Fig. S1A-D). In addition, no sample or patient group-specific misalignment of cell clusters was observed (Fig. 1B, Fig. S1E), thus no batch correction of the dataset was needed. Cell-level cell-type annotation was mainly performed with the reference-based SingleR method (v1.10.0) (*25*) using the Monaco immune data as a reference (*26*). The most frequent cell-level annotation was assigned to each cluster, and cell clusters with the same annotation were readily combined. We then confirmed and refined the cluster-level annotations manually using cluster markers detected from the scRNA-seq and CITE-seq data (Fig. 1D-E) and annotated additional cell types of interest (such as GC B cells, Tfh-like, GC Tfh, Tfr) that were missing in the reference data for SingleR. We also performed sub-clustering of B cell subsets, for which cluster annotations were performed manually based on cluster markers detected from the scRNA-seq and CITE-seq data (Fig. 2, Data file S6,8,10).

We then used the cell clusters as defined by the annotation above for further analyses. To identify differentially abundant cell types in MS, cell type composition was evaluated between MS and control samples by comparing the proportion of the cell types in the two conditions. Differential gene expression was identified for each cell type using the pseudobulk approach, which has been reported to outperform other single-cell DE methods (*85*), by sum aggregating the RNA counts of genes per cell type per sample, and comparing the pseudobulk gene expression within each cell type between MS and controls using DESeq2 (v1.36.0) (*86*). Biological pathway analysis was then performed for each cell type using the pathway-express method (R package ROntoTools v2.24.0) (*87*, *88*).

Basic evaluation and comparison of immune single-cell level TCR and BCR repertoire clonality/diversity and clonal overlap were done with scRepertoire (v1.6.0) (*89*) and in-house custom R script. We scanned the VDJdb database (release 2022-03-30) (*90*) of known TCR-epitope targets for possible specificity of our TCRs using tcrdist3 (v0.2.2) (*91*), with a tcrdist3 distance of less or equal to 12 per alpha and beta chain (translating to one amino acid mismatch per chain) used to assign possible targets for our TCRs. This was done for paired TCRab, and the specificity profiles were compared between MS patients and controls. For validation, TCRGP (1.0.0) (*63*) models were made for the 28 HLA class I-restricted epitopes from endemic viruses (CMV, EBV, Influenza A). For the predictions, only CD8+ T cells were used and a threshold corresponding to a false positive rate (FPR) of 5% was determined for each epitope separately from the receiver operating characteristic (ROC) curves obtained from the cross-validation experiments, as described previously. For BCRs, clonality definition in scRepertoire was used, i.e., BCRs using the same V gene for both heavy and light chains (HC and LC), and that have >85% nucleotide similarity in the HC and LC CDR3s (with normalized Levenshtein distance) were grouped into one clonotype. For comparison with our results, we also used the clonotype definition by Lanz et al. 2022 (*8*), i.e., sharing the same HC and LC V and J genes, and exhibiting >70% amino acid identity within the HC and LC CDR3s (with normalized Levenshtein distance).

Human leukocyte antigen (HLA) genotypes were predicted from the scRNA-seq data with arcasHLA (*92*), which has recently been shown to have better accuracy than other similar tools (*93*). The HLA-DR15 allele with the strongest risk for MS, HLA-DRB1*15:01(*74*), was present in all (3/3) of the healthy controls and half (3/6) of the MS patients (Data file S1), suggesting that the observed TCR/BCR repertoire changes in our MS patients are more likely caused by other reasons than due to HLA-DR15 allele.

### Intercellular communication analysis

We used the NicheNet package (v. 2.0.1) (*48*) to study differences in intercellular communications between observed cell types. In each combination of sender and receiver cell type, genes that were expressed in at least 10% of the sender/receiver cell type population and present in the pre-built NicheNet prior ligand-receptor interaction model were chosen as potential receptor and ligand genes. In addition to ligand activity analysis, the ligands were prioritized based on condition-specificities of the ligands and receptors across all cell types to highlight any ligands and receptors expressed differentially between MS patients and controls. We visualized the top ligand-receptor pairs for each sender-receiver cell type combination using the mushroom plot function offered by NicheNet. In addition, the MultiNicheNet package (v. 1.0.3) (*52*) was used to observe differences between controls and MS patients. For the analysis, we followed the main steps in the MultiNicheNet vignette.

### Detection of viruses from scRNAseq data

To detect any EBV-producing cells, we used the Vulture pipeline (*62*), which utilizes viral and prokaryote genome files from the ViruSITE (*60*) and NCBI (*61*) databases along with alignment tools to map 10x scRNA-seq reads to microbial genomes. Vulture was used together with the CellRanger alignment tool to map the raw scRNA-seq data to the microbial genomes. After performing QC on the mapped reads, we normalized the obtained counts across each sample. This enabled comparing the counts of microbial features across samples.

### Hybrid-capture sequencing of DNA viruses from dcLN FNAs

The DNA was extracted using the QIAamp DNA Mini Kit (Qiagen), following the manufactureŕs protocol for tissue extraction. The extracts were eluted in 60 ul of AE buffer and the DNA was mechanically fragmented with a Covaris E220 with a target length of 200 nt. Subsequently, the libraries were prepared with the KAPA HyperPlus kit (Roche) using unique Dual Index Adapters (Roche). Targeted enrichment of the viral DNAs was performed using a custom panel of biotinylated RNA probes (Arbor Biosciences) as described by Toppinen et al (*22*). Each sample was individually enriched via two rounds of hybridization, following the manufacturer’s recommendations for low-input DNA (MyBaits v5 kit; Arbor Biosciences). The probes were 100 bp in length and designed with 2X tiling. Kapa Universal Blockers (Roche) were used to block unspecific binding to the adapters during hybridization.

During library preparation and viral enrichment, the libraries were amplified 3x13-25 cycles. The clean-up steps were performed with 1x KAPA HyperPure Beads (Roche). The enriched libraries were quantified with the KAPA Library Quantification Kit (Roche) using Stratagene 3005P qPCR System (Agilent) and pooled for sequencing on NovaSeq 6000 (one lane, S4, PE151 kit; Illumina).

### Data analysis of hybrid-capture sequencing

The data analysis was done with TRACESPipeLite, a streamlined version of TRACESPipe (*94*). The paired-end reads were trimmed and collapsed with AdapterRemoval, cutting ambiguous bases at the 5’/3’ termini with quality scores below or equal to two. Reads shorter than 20 bases were discarded. FALCON-meta was used to find the highest similar reference from the NCBI viral database (*95*). The reads were aligned with BWA (*96*) using a seed length of 1000 and a maximum diff of 0.01. Read duplicates were removed with SAMtools (*97*) and the consensus sequences were reconstructed with BCFtools (*98*). The coverage profiles were created with BEDtools (*99*). When in low breadth coverage (< 10%), the individual reads were manually inspected and confirmed by BLAST. The pipeline is freely available, under the MIT license, at https://github.com/viromelab/TRACESPipeLite, along with the code (included in the TRACESPipeLite repository).

### Quantitative multiplex PCR (qPCR) of saliva

DNA extraction was optimized by adapting the protocol from Qiagen QIAamp DNA Blood Mini Kit and Qiagen DNeasy Blood & Tissue Kit (Qiagen, Hilden, Germany) to get the highest yield of DNA. Initially, 8 ml of phosphate-buffered saline (PBS) (Lonza, Basel, Switzerland) was added to 2 ml of each saliva sample. The samples were then centrifuged at 1800 g for 10 minutes. The pellet was resuspended in 180 µl ATL buffer by pipetting up and down. The sample was pipetted into a new tube with 40 µl proteinase K, vortexed, and incubated at 56°C with shaking for 1 h. After this, 200 µl AL buffer was added followed by vortex and incubation at 70°C for 10s min. From here, the protocol continued following the Qiagen QIAamp DNA Blood Mini Kit Spin Protocol. In the last step, DNA was eluted into 60 µl AE buffer with an incubation time of 5 min. The samples were stored at -20°C until further processing.

Multiplex qPCR method HERQ-9 was used to detect and quantify nine HHVs simultaneously from the saliva samples as described earlier by Pyöriä et al (*100*). Briefly, HERQ-9 is designed in three triplex-qPCR reactions, the first one amplifying herpes simplex viruses 1 and 2 (HSV-1 and 2) and varicella-zoster virus (VZV), the second one amplifying EBV, human cytomegalovirus (HCMV), and Kaposi’s sarcoma-associated herpesvirus (KSHV), and the third one amplifying HHV-6A, -6B, and -7. The viral loads were normalized per 10^6^ cells, determined with the human single-copy gene RNase P qPCR (*101*).

### Flow cytometry

For cell surface staining, cells were suspended in an antibody cocktail containing LIVE/DEAD® Fixable Green Dead Cell Stain (ThermoFisher), which was prepared using Brilliant Stain Buffer (BD Biosciences). The cell suspension was then incubated at +4°C for 30 minutes and subsequently washed twice with FACS staining buffer (PBS, pH 7.4, 10% FBS, and 2 mM EDTA). Flow cytometry data were acquired with LSR Fortessa (BD Biosciences) and analyzed with FlowJo (BD Biosciences, LLC). Detailed information about the antibodies used in this study can be found in Data file S13.

### Statistics

Either the t-test or evaluation of the standardized Pearson residuals using the Chi-Square test was used to compare two different categories. In the chi-squared test, all cells were pooled by subject group and the overall cell type distribution frequency of the standardized Pearson residuals was compared between MS patients and healthy controls. Euclidean dissimilarity distances and either Ward’s or complete agglomerative clustering were used in the correlation plots. In differentially expressing markers (both scRNA-seq and CITE-seq) analysis, the non-parametric Wilcoxon rank sum test was used and adjustments for multiple testing were performed with Bonferroni correction.

Nominal and adjusted p-values (Padj) of less than 0.05 were considered significant. In the box plots, the vertical line corresponds to the median, the box corresponds to the interquartile range (IQR), and the whiskers are 1.5 × the IQR. All calculations were done with R (4.2.1, 4.3.1) or Python (3.7.4).

## Supplementary Materials

Supplementary Material on cluster phenotyping Supplementary Figures 1-5

Description of supplementary datafiles

## Data Availability

The Seurat-objects of the processed scRNA-sequencing data including TCR and BCR and CITE-seq data are available at Zenodo (10.5281/zenodo.10006020) with restricted access due to GDPR regulations and data can be accessed by placing a request via Zenodo. Differentially expressed gene sets are available in supplementary data files. The reconstructed near-complete HHV-7 genomes are available at GenBank (OR634979 and OR634980).

https://doi.org/10.5281/zenodo.10006020

## Acknowledgments

We are grateful for the patients and healthy controls for their participation, and without whom this study would not have been possible. We thank Marjo Rissanen and Kalla- Maria Tiilikainen for their help with sample processing. The authors would like to thank the FIMM Single-Cell Analytics unit supported by HiLIFE and Biocenter Finland for scRNA and CITE-seq services.

## Funding

Academy of Finland grant 332186 (SML)

Competitive Research Funding of the State of Finland governed through Helsinki University Hospital grant TYH2020317 (SML)

Finska Läkaresällskapet (SML) Suomen MS-Säätiö (JS)

Emil Aaltonen Foundation (JS) Instrumentarium Science Foundation (JS) Suomen Lääketieteen säätiö grant 6006 (MP) Liv och Hälsa (MP)

## Author contributions

Conceptualization: SML, JS, MP, EK, PJT

Methodology: JS, DY, SML

Software: DY, JH

Validation: JS, DY, MP

Formal Analysis: JS, DY, NK, PD, JH, MP

Investigation: JS, PD, KN, GK, RL, MP, SML

Resources: SML, MS, EK, MP

Data Curation: JS, DY, NK, PD, KN, MP

Writing – Original Draft: JS, DY, NK, SML

Writing – Review & Editing: SML, JS, DY, JH, KN, PJT, EK, MP

Visualization: JS, DY, NK, JH

Supervision: EK, SML

Project administration: JS, SML

Funding acquisition: SML, EK, MP

## Data and materials availability

The Seurat-objects of the processed scRNA-sequencing data including TCR and BCR and CITE-seq data are available at Zenodo (10.5281/zenodo.10006020) with restricted access due to GDPR regulations and data can be accessed by placing a request via Zenodo. Differentially expressed gene sets are available in supplementary data files.

The reconstructed near-complete HHV-7 genomes are available at GenBank (OR634979 and OR634980). The code to reproduce the key findings is available on GitHub (https://github.com/SarkkinenJ/cervical_lymphnodes_MS.git).

