## Supplementary material and figures for "Deep cervical lymph nodes of patients with multiple sclerosis show dysregulated B cells in the presence of Epstein-Barr virus"

#### Table of Contents


- Supplementary Material on cluster phenotyping
- Supplementary Figures
- Description of supplementary datafiles

#### Supplementary material on cluster phenotyping

We performed cell type determination using a combination of automatic annotation with SingleR(*25*) using the Monaco reference(*26*) and manual annotation aided by the CITE-seq data that we had for 4/6 MS patients and all (3/3) healthy controls (Fig. 1C). Manual cell type determination was used especially for T and B cell subsets using a combination of key markers on both transcript and protein levels (Fig. 1D-E, Data file S2-3).

Clusters of T and B cell subsets were identified using combination of key markers on both transcript and epitope level (Fig. 1D-E, Data file S2-3). The naïve CD4+ T (naïve CD4), characterized with CD45RA upregulation in CITE-seq data and absence of *FAS*, were the most prevalent of all cells (≈ 40%, Fig. 1F). Using CD45RO protein expression and absence of CD45RA, we identified activated effector CD4+ T cell cluster (Effector CD4), which was absent for CD45RA, and a CD4+ T cell cluster with intermediate CD45RA and CD45RO expression (Intermediate CD4), which also expressed activation markers such as *ICOS* and *CD27*. Tfh cells (Tfh-like, GC Tfh) also upregulated CD45RO and were identified by canonical *CXCR5* expression(*27*) supported with *CD40LG*(*49*) and represented 11% and 0.5 % of all cells. The GC Tfh cluster expressed *BCL6*(*28*), *ICOS*(*29*), *TOX* and *TOX2*(*30*), and *PDCD1*(*31*) and likely a more active phenotype of Tfh (here GC Tfh) cells since they also expressed *SH2D1A* (*SAP*)(*32*), and *IL21*(*33*) abundantly as a sign of increased interaction with GC B cells (Fig. 1E). Regulatory T (Treg) and Tfr cell clusters were characterized with upregulation of *FOXP3*, *IKZF2*, and *CTLA4*(*19*), of which the Tfr cluster differently expressed ICOS(*34*) and PDCD1 on RNA and protein level (Fig. 1D-E, Data file S2-3). As expected, CD4+ T cells were more common than CD8+ T cells (64% vs 15% of all cells), which clustered into two clusters: a naïve cluster representing 2/3 of all CD8+ T cells characterized with CD45RA, and a memory cluster expressing CD45RO, which represented 1/3 of CD8+ T cells.

The B cell compartment, characterized with *CD79A* and *CD19* expression compared to non-B cells, clustered into four main clusters (Fig. 1C): naïve B cells differentially expressed *IGHD* and *IGHM* in the absence of *IGHG* and *CD27*, whereas memory B (MB), plasma cells / plasma blasts (PC/PB), and GC B cells differentially expressed both *CD27* and *IGHG*, in addition to *Notch2* in MB, *PRDM1* and *SDC1* in PC/PB, and *BCL6* and *AICDA* in GC B cell cluster (Fig. 1D-E, Data file S2-3) (*46,47*).

### Supplementary Figure 1.

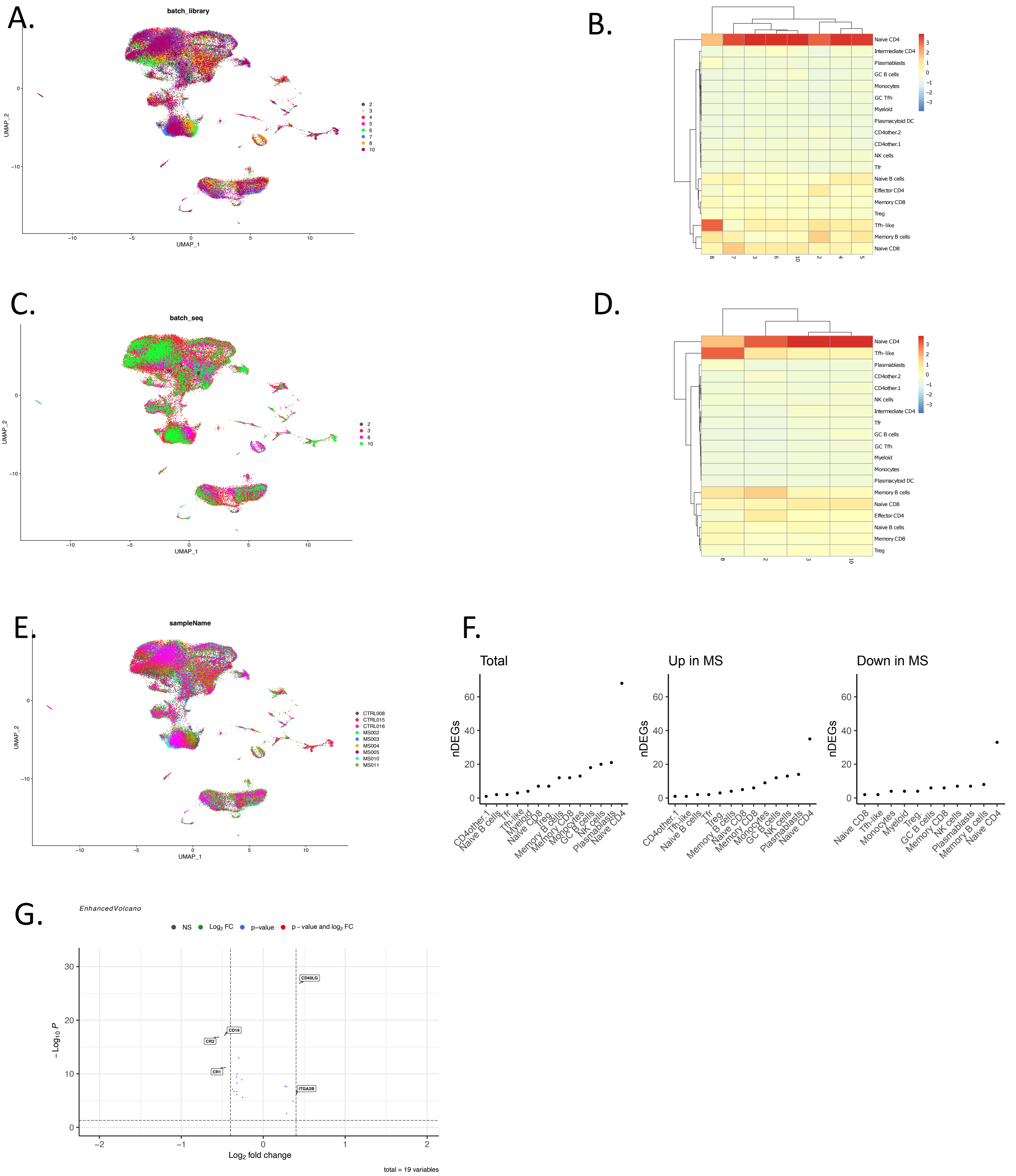


**Supplementary Figure 1: Cell clusters of cLNs and DEGs in MS patients compared to controls**
A) Uniform Manifold Approximation and Projection (UMAP) of identified dcLN scRNA-seq cell clusters between library preparation batches. B) Heatmap with dendrograms show similar cell proportions in each library preparation batches. C) UMAP) of identified dcLN scRNA-seq cell clusters between sequencing batches. D) Heatmap with dendrograms show similar cell proportions in each sequencing batches. In A-D), labels reflect the batch number for library preparation and sequencing. Further information of library preparation and sequencing batches in samples is shown in Supplementary Table 1A. In B) and D), Euclidean distance and complete linkage was used for agglomerative hierarchical clustering. E) UMAP) of identified dcLN scRNA-seq cell clusters between separate samples. F) The number of differentially expressing (DE) genes of pseudobulked cel types in MS patients compared to healthy controls in each cell type. G) Volcano plot of DE proteins in GC B cells in MS patients compared to healthy controls. DE proteins were calculated from CITE-seq data using single cell DE. DE proteins with adjusted p value $< 0.05$ are shown as dots and with log2FC $> 0.4$ or $< -0.4$ with labels.

### Supplementary Figure 2.

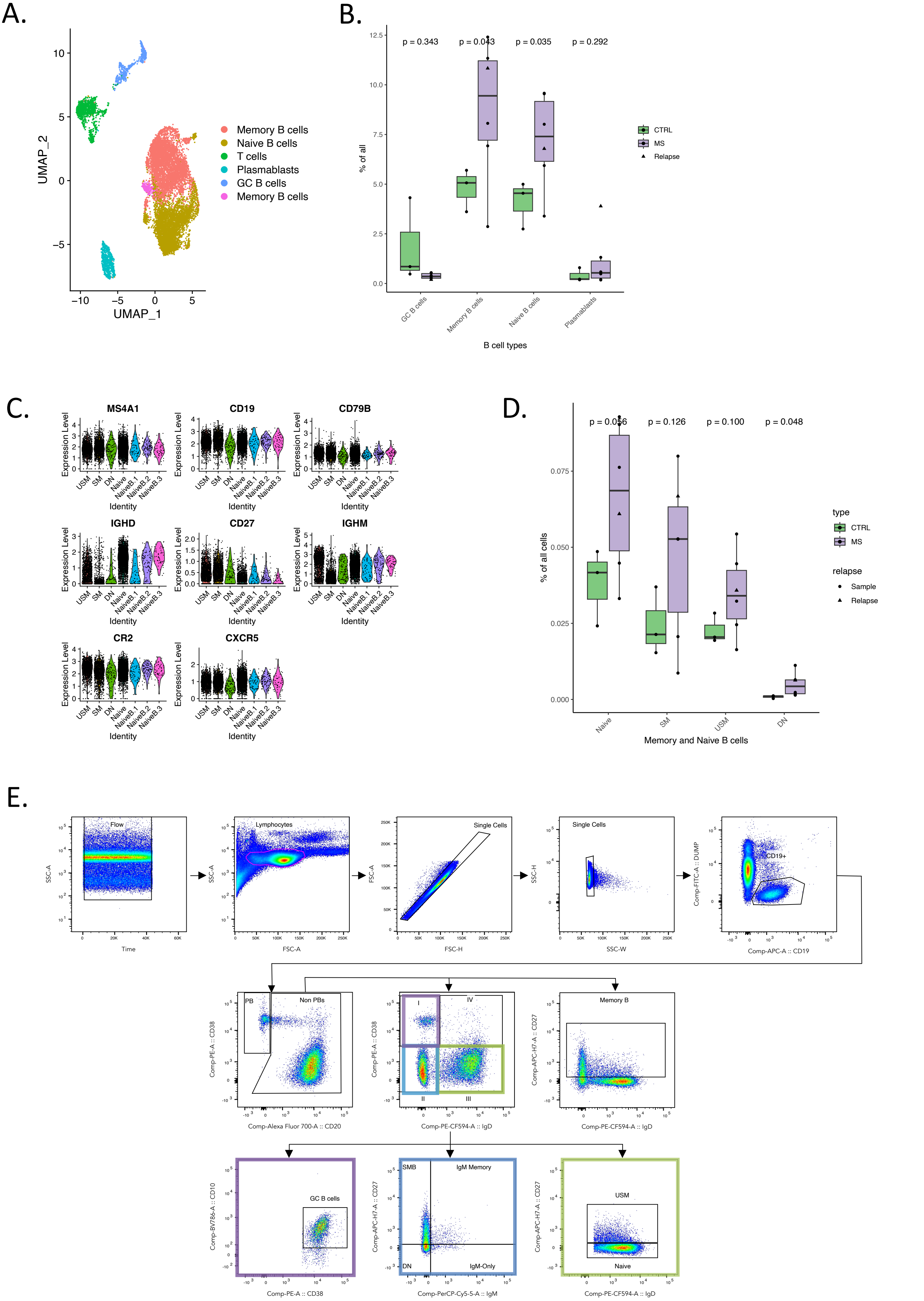

F.

Flow cytometry

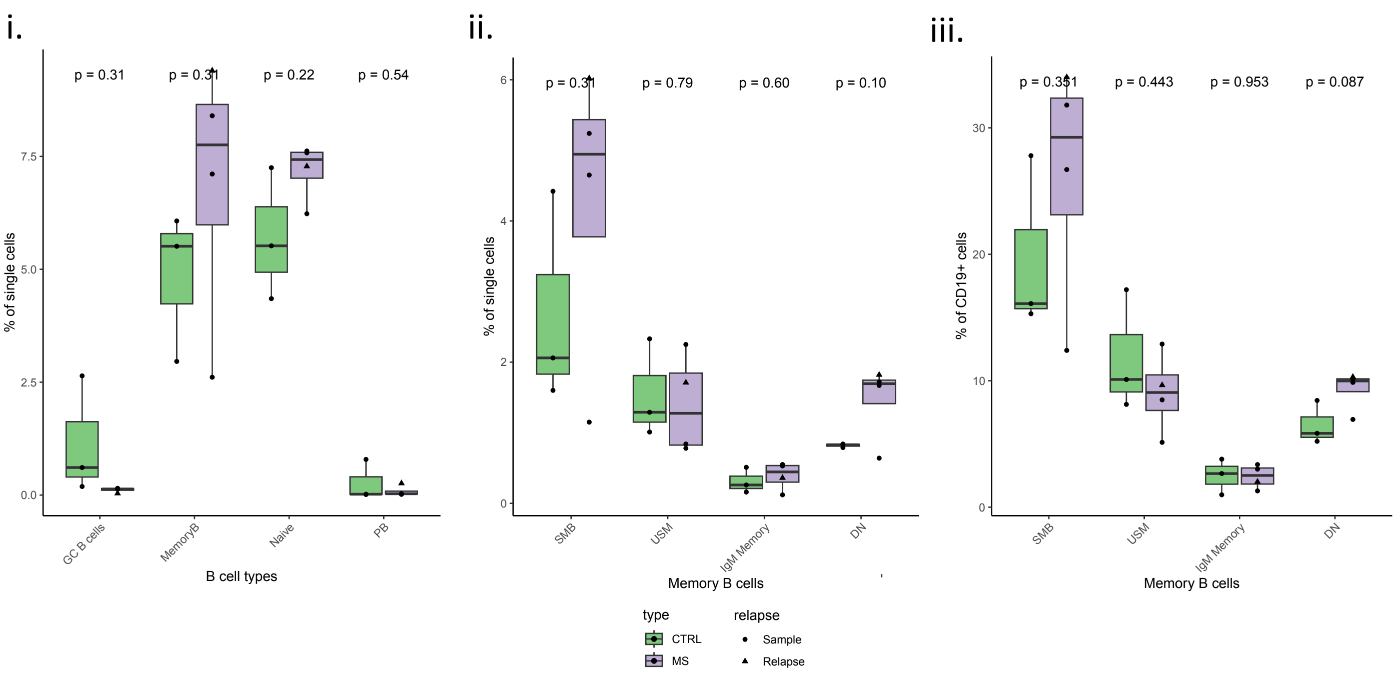


G.

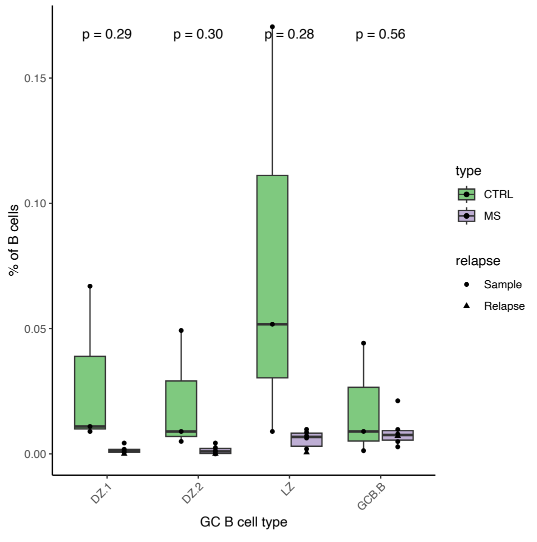


**Supplementary Figure 2: Further clustering of B cells**
A) UMAP presenting re-clustered plasmablasts, and naïve, memory, and GC B cells identified in Figure 1C, E-F. One cluster contained T cells and was filtered away from the further analysis of B cells. B) Box plot of B cell subsets (presented in A) between MS patients and controls. C) Expression of key CITE-seq markers in identified memory and naïve B cell subsets presented with violin plots. D) Box plot of memory and naïve B cell subsets (% of all cells) between MS patients and controls. E-F) B cell subsets in dcLNs of MS patients (n = 4) and healthy controls (n = 3) using flow cytometry. The gating strategy (E) is shown for healthy control and was adopted from Sanz et al (42) to identify B cell populations from dcLNs. Dump channel included CD3, CD14, and a dead cell marker. F) Box plots of proportions of main (i) and memory (ii-iii) B cell subsets. Marker combinations to define subsets: GC B cells, CD19+CD20+CD38++CD10+IgD-; Memory B cells, CD19+CD20+CD27+; Naïve B cells, CD19+CD20+IgD+CD27-CD38-; PBs, CD19+CD38+CD20-; SMB, CD19+CD20+CD27+IgD-IgM-CD38-; USM, CD19+CD20+CD27+IgD+IgM-CD38-; IgM Memory, CD19+CD20+CD27+IgM+IgD-CD38-; DN, CD19+CD20+CD27-IgD-IgM-CD38-. G) Box plot of GC B cell subsets (% of all B cells) between MS patients and controls from scRNAseq data. In B, D, F-G), an unpaired t-test was used, and the patient with an active relapse is highlighted with a triangle.

### Supplementary Figure 3.

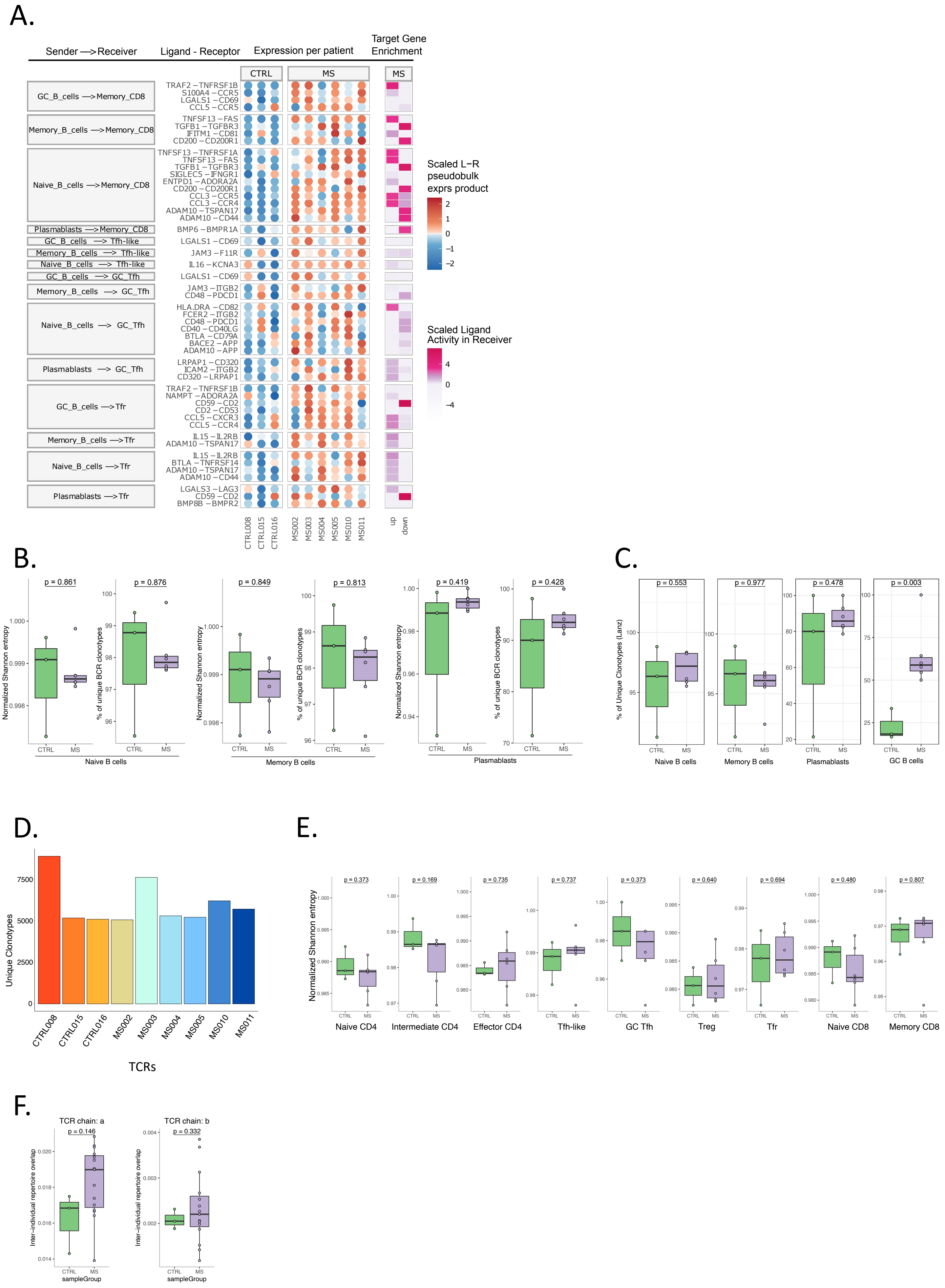

G.

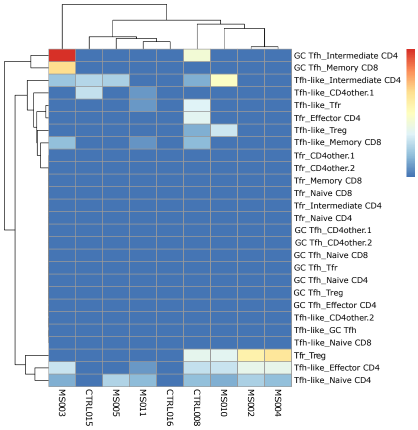


H.

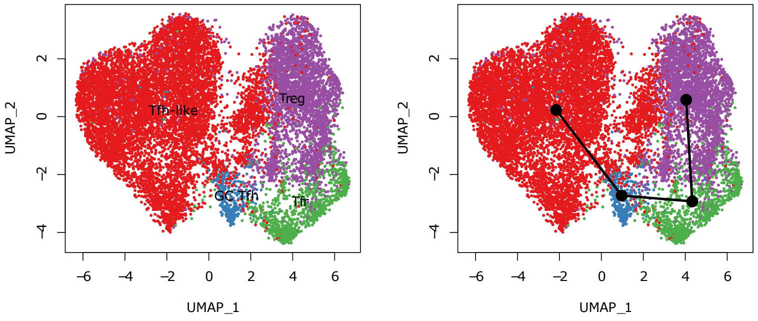


I.

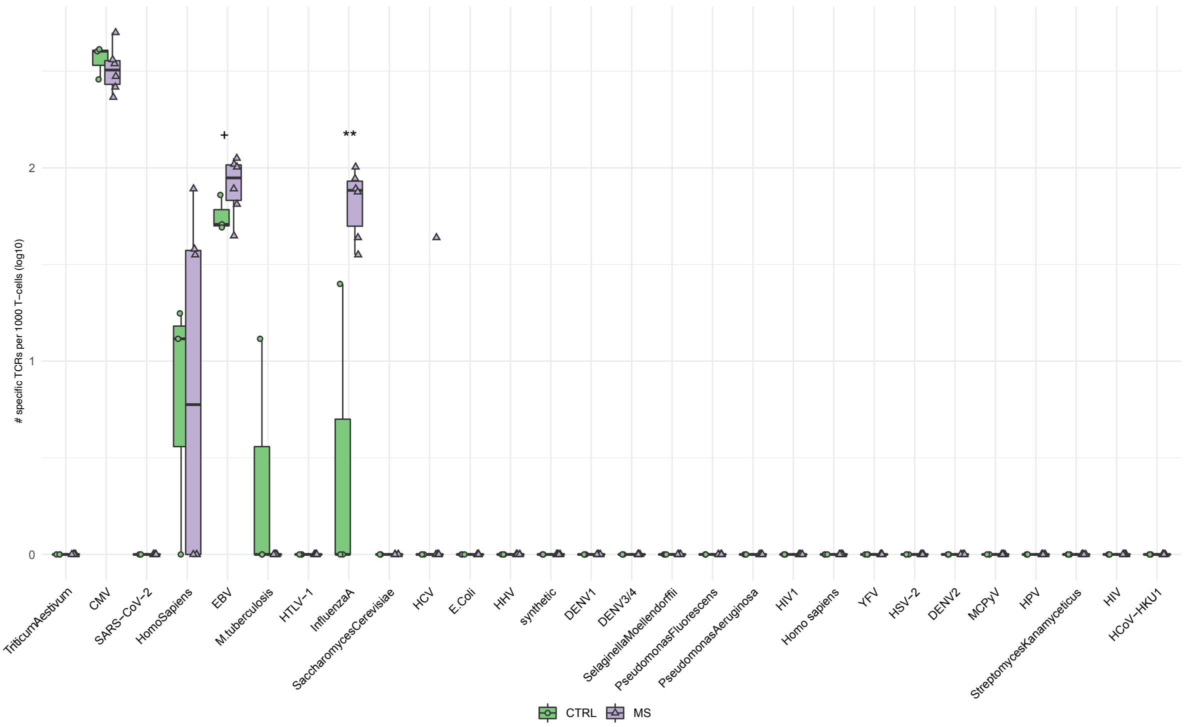


**Supplementary Figure 3: Intercellular communication, TCR, and BCR analysis**

A) Summary heatmap of intercellular communications with B and T cell subsets in MS patients compared to healthy controls using MultiNicheNet. Pseudobulk expression of predicted ligand–receptor pairs in sender and receiver cells in each dcLN sample are shown in dot plots, where red colour corresponds to increased and blue to decreased ligand-receptor pair. The summarized scaled (z-score) NicheNet ligand activity in receiver cell type in MS patients´ dcLNs compared to healthy controls is shown on the right. B) BCR diversity of naïve, memory B cells, and plasmablasts. For each B cell subset, the left panel presents the normalized Shannon entropy (diversity), and the right panel percentage of unique clones in each sample. C) BCR diversity assessed by using the definition by Lanz et al. 2022 (see Methods and References). For B-C), an unpaired t-test was used to compare diversity estimates between MS patient and control repertoires. D) Bar plot illustrates the number of unique TCR clonotypes in each sample. E) Box plot of TCR diversity in T cell subsets using Shannon entropy. F) Box plots of inter-individual repertoire overlap of TCR alpha (left) and beta (right) amino acid chains between MS patients and controls. TCR overlap rates for every possible pair between patients and controls were compared to evaluate any convergent selection of functional TCRs in patients. In E-F, an unpaired t-test was used. G) Nucleotide level paired TCR overlap rate between T cell subsets and Tfh-like, GC Tfh, and Tfr cells, patients vs. controls was evaluated to assess clonal lineage across T-cell subsets. The overlap rate was defined as multiplying the number of overlapping clonotypes between two subsets by two divided total number of clonotypes in both subsets. Clustering was performed with Euclidean distance and complete linkage method. H) UMAP of reclustered Tfh-like, GC Tfh, Tfr, and Treg cells on left, and predicted lineage trajectory plot using the Slingshot package. I) Box plot of predicted antigen specificity of GC Tfh cell clonotypes between MS patients and controls (+, an unpaired t-test p-value = 0.08, log2FC = 0.56; **, p-value = 0.003, log2FC = 3). The VDJdb database was scanned for TCRab clonotypes using tcrdist3.

### Supplementary Figure 4.

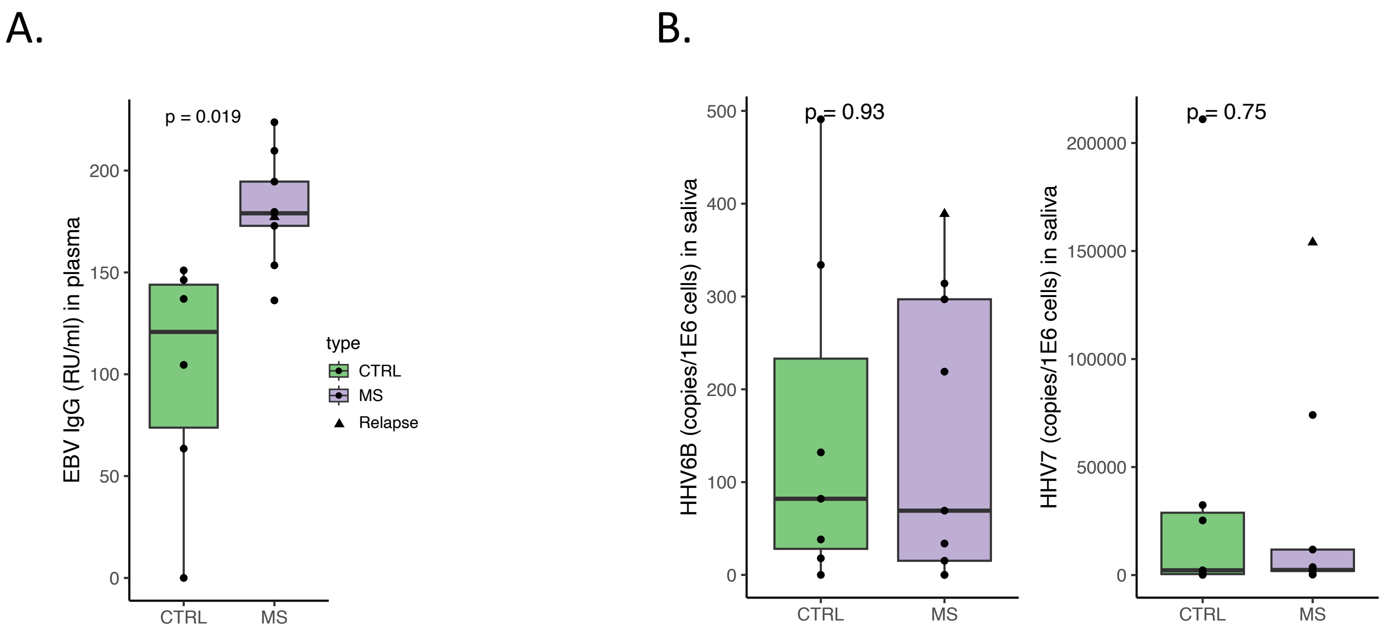


**Supplementary Figure 4**

A) Box plot of EBV viral capsid antigen (VCA) titer between MS patients and controls using enzyme-linked immunosorbent assay (ELISA). B) Copy numbers of HHV6B (left) and HHV7 (right) per million cells between MS patients and controls are presented with box plots. The patient with a relapse is highlighted with a triangle. In A-B), an unpaired t-test was used to compare MS patients and controls.

### Supplementary Figure 5.

A.

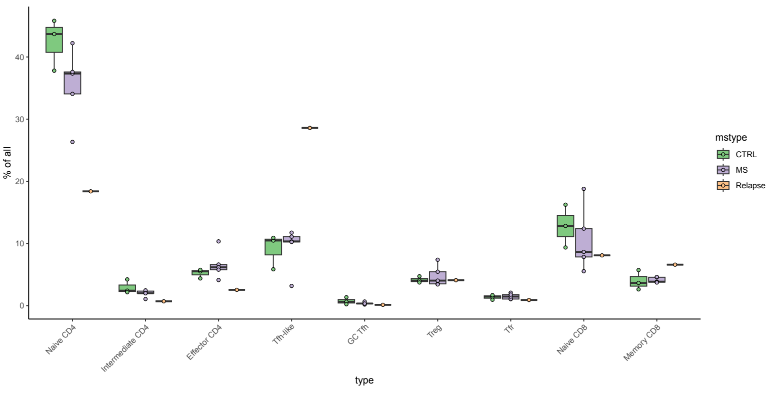


B.

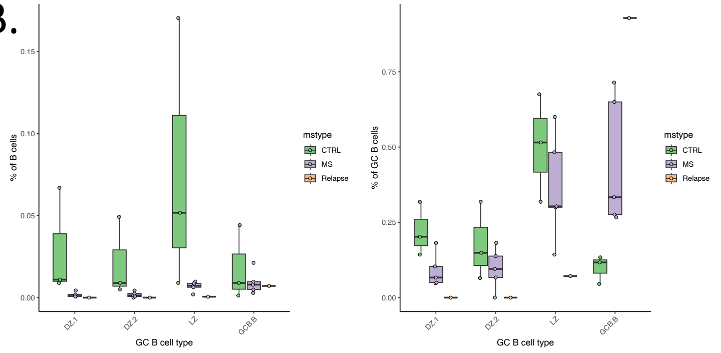


C.

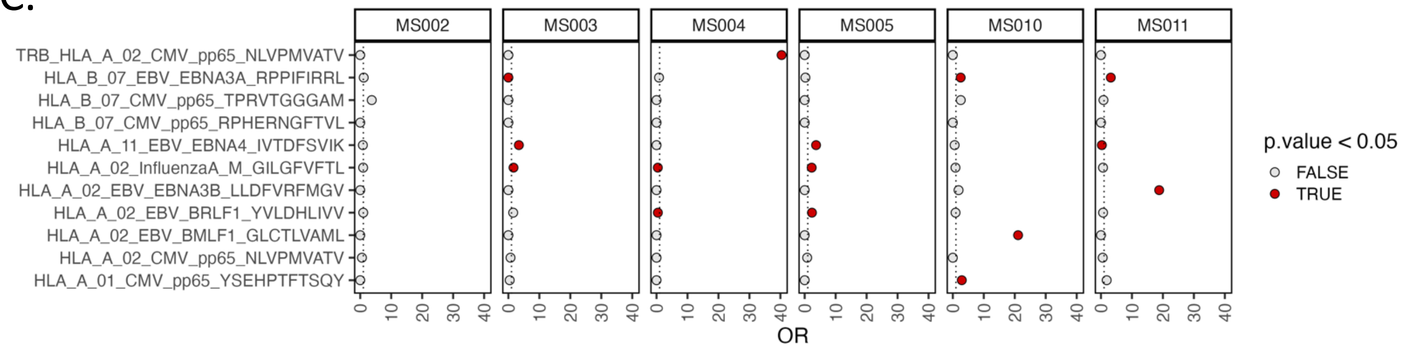


**Supplementary Figure 5: MS patient with an active relapse has altered number of Tfh cells and decreased GC B cells**

A) Box plots of proportions of T cell subsets in MS patient with relapse (orange) compared to other MS patients and controls. B) The proportion of GC B cell subsets of all B cells (left) and all GC B cells (right) presented with box plots. C) Odds ratios of TCR predictions for expanded CD8+ T cell clones (TCR clone ≥ 2 times in a sample) targeting viral antigens in individual MS patients compared to healthy controls using TCRGP. The median predictions of expanded CD8+ T cells for each viral antigen in healthy controls is highlighted with a dotted line.

### Description of supplementary datafiles

File Name: supplementarytable.xlsx

Description:

1. Patient details (sheet: 1.clinical_info)
2. scRNAseq markers used for annotation of main clusters (sheet: 2.mainsubsets_genes)
3. scCITE-seq markers used for cluster annotation (sheet: 3.mainsubsets_CITEseq)
4. Cell counts of main cell subsets (obtained from 2) (sheet: 4.cellcounts_mainsubsets)
5. Differently expressed (scRNAseq) genes in main cell subsets (obtained from 2) between MS patients and healthy controls (sheet: 5.DE_genes_MSvshc_mainsubsets)
6. scRNAseq markers used for cluster annotation of B cells obtained from 3) (sheet: 6.subclustBcells_genes)
7. Cell counts of B cell subsets (sheet: 7.cellcounts_B_cells)
8. scRNAseq markers used for cluster annotation of naïve and memory B cells obtained from 6) (sheet: 8.subclust_MemNaiveB_genes)
9. Cell counts of Memory and Naïve B cell subsets (sheet: cellcounts_MemNaiveB)
10. scRNAseq markers used for cluster annotation of GC B cells obtained from 6 (sheet: 10.subclust_GCB_genes)
11. Cell counts of GC B cell subsets (sheet: 11.cellcounts_GCB)
12. Top 20 pathways (Go biological processes) in GC B cell subsets defining genes (p.adj < 0.05) (sheet: 12.pathway_GCB)
13. Flow cytometry antibodies (sheet: 13.flow_cytometry_antibodies)
